# Epigenetic signatures of regional tau pathology and cognition in the aging and pathological brain

**DOI:** 10.1101/2024.11.07.24316933

**Authors:** David C. Goldberg, Anil R. Wadhwani, Nadia Dehghani, Lasya P. Sreepada, Hongxiang Fu, Philip L. De Jager, David A. Bennett, David A. Wolk, Edward B. Lee, Charles L. White, Jamie M. Walker, Timothy E. Richardson, PART Working Group, Kurt Farrell, John F. Crary, Wanding Zhou, Corey T. McMillan

## Abstract

Primary age-related tauopathy (PART) and Alzheimer’s disease (AD) share hippocampal phospho-tau (p-tau) pathology but differ in ß-amyloid burden and degree of p-tau severity and spread. Thus, PART provides a human model to understand the mechanisms of age and amyloid-independent modifiers of p-tau. Given the dynamics of DNA methylation over the lifespan, we (1) performed an epigenome-wide association study of PART that nominated 13 loci associated with p-tau; (2) developed two novel epigenetic clocks predictive of p-tau in age-, and ß-amyloid-independent manners: “TauSeverity” relates hippocampal p-tau severity in PART and AD and synaptic transmission genes; “TauSpread” relates to p-tau spread to frontal cortex of AD and neuroinflammatory genes; and (3) a machine learning classifier that identifies low- and high resilience individuals with overlapping neuropathological features but distinct epigenetic, transcriptomic, and clinical features. We conclude that the epigenome contributes to the severity and spread of p-tau pathology, guided by distinct pathways with cognitive consequences.

## INTRODUCTION

Neurofibrillary tangles (NFTs), consisting of hyperphosphorylated tau (p-tau), are a ubiquitous feature of aging present in nearly every human brain older than 50 years of age^1–5^. However, the overall p-tau burden, as well as the associated degree of cognitive impairment, vary widely across individuals, even after accounting for age and co-pathology^6,7^. Frequently, p-tau burden is classified using Braak staging, which describes a stereotypical progression of spread from the medial temporal lobe to the associative neocortex^8^. However, this framework does not fully capture inter-individual variability in p-tau severity within a region. Emerging evidence suggests that these two distinct dimensions of p-tau pathology – severity and spread – may better explain its heterogeneity.^9–11^ Severity reflects the local burden of p-tau aggregates within a brain region, whereas spread describes the extent to which p-tau propagates beyond its initial deposition sites.^12,13^

Two related conditions, Alzheimer’s disease (AD) and Primary Age-Related Tauopathy (PART), represent two ends of the spectrum of p-tau pathology, which varies in both severity and spread. P-tau in both conditions is molecularly similar^6^, and genetic risk factors partially overlap^14^. Both exhibit variable age-associated hippocampal p-tau severity, yet they diverge in neocortical p-tau spread^6,15–21^. In PART, p-tau remains relatively confined to limbic structures, whereas in AD, it spreads extensively, first to adjacent temporal and association cortices and later to more distant midfrontal cortex^2,8^. Another key distinction is the presence of ß-amyloid co-pathology in AD but not in PART. As a result, PART serves as a human model to investigate ß-amyloid-independent modifiers of p-tau severity, and comparing PART and AD enables the dissection of both ß-amyloid-independent and -dependent mechanisms underlying p-tau spread. While some have suggested that PART may represent an early stage of AD^22^, distinct genetic and anatomical features suggest otherwise. The frequency of AD risk alleles differ between AD and PART^14^, a novel locus in *JADE1* has been implicated in PART-specific risk^23^, and PART preferentially involves the CA2 hippocampal subfield, which is largely spared in AD^24^.

Given the genetic and anatomical differences between PART and AD, the two conditions appear to be distinct trajectories of brain aging. Because DNA methylation (DNAm) changes widely across the lifespan, we hypothesized that distinct epigenetic programs also underlie p-tau severity and spread. While DNAm patterns have been characterized in AD^25–30^, prior studies have been limited in several ways. First, blood-derived DNAm epigenetic clocks have linked accelerated epigenetic aging to AD severity^31,32^, but the biological basis of these clocks remain obscure and lack brain-specific associations^33^. Second, while studies of cortical brain DNAm have identified CpGs associated with AD neuropathologic change, such as Braak stage^26^ and senile plaques^25^, they have been performed without regard to within-region severity or the complex interaction of p-tau and ß-amyloid. Therefore, it remains unclear whether these epigenetic changes are a feature of normal brain aging or if they are a consequence of AD pathophysiology.

To understand the epigenetic contributions to p-tau in PART and AD we report several computational approaches to analyze frontal cortex DNAm to disentangle associations between p-tau, ß-amyloid, and aging across the pathological spectrum of PART and AD. We conducted the first epigenome-wide association (EWAS) study of PART to identify amyloid-independent modifiers of p-tau severity. We then generated two novel epigenetic clocks – “TauSeverity” and “TauSpread” – that are predictive of the respective distinct dimensions of age-independent p-tau burden. Lastly, we developed a comparative multivariate classifier of PART and AD that distills the shared biological programs contributing to both conditions from those uniquely protective in PART and identifies an epigenetic signature associated with cognitive resilience.

## RESULTS

### Description of cohorts

We generated frontal cortex DNA methylation profiles for 260 individuals in the PART working group (PWG) cohort comprised of autopsy-confirmed PART cases (Braak NFT stage I-IV, CERAD = None) with digital histology measures of p-tau (NFT) density in AT8-stained tissue sections of hippocampus and frontal cortex, as previously reported^34^. All PWG samples were interrogated using the Illumina EPIC Human Methylation array for ∼850K CpGs (Methods: DNAm data generation). We also performed secondary analyses using dorsolateral prefrontal cortical data available from 707 individuals from the Religious Orders Study and the Rush Memory and Aging Project (ROSMAP) cohort^25^, which includes 176 cases with PART. Cohort characteristics are summarized in **Supplemental Table 1**. All ROSMAP samples were interrogated for DNAm using the Illumina Human Methylation 450 beadchip array and a subset had matched gene expression data generated by the Illumina TruSeq method with modifications as previously described^35^. The unsupervised clustering of DNA methylomes for each cohort was primarily driven by sex and sample plate and did not reveal relationships to hippocampal p-tau load, Braak stage, or CERAD score (**Extended Data Fig. 1a**). To assess for potential confounding due to cell subtype variation, we employed a reference atlas for 20 major cellular subtypes in brain obtained from publicly availably single cell whole genome bisulfite sequencing data^36–38^. We detected a slight enrichment of superficial cortical layer cell types in the ROSMAP cohort relative to the PWG cohort (**Extended Data Fig. 1b,c**). Nonetheless, cell-type proportion principal components and individual cell-type proportions did not associate with hippocampal p-tau severity in either cohort (**Extended Data Fig. 1d,e**), suggesting that downstream analyses were not confounded by cell type composition.

### An EWAS and a novel epigenetic clock, “TauSeverity”, capture hippocampal tau variation in PART

To identify epigenetic contributions to the amyloid-independent inter-individual variation in hippocampal p-tau severity, we performed an EWAS in the PWG cohort (N=260 PART), adjusting for age, sex, and sample plate. We identified 2 CpG sites associated with hippocampal p-tau (*cg04905912* and *cg17649772,* Bonferroni-corrected significance threshold < 0.05) and an additional 11 CpG sites that met a less stringent False Discovery Rate (FDR) < 0.05 (**Fig. 1a**, **Fig. 1b, and Extended Data Fig. 2a**). To our knowledge, methylation variation at these 13 loci has not been previously associated with either amyloid or p-tau pathology in aging or AD in prior brain methylation studies^25–30^.

**Fig. 1:**
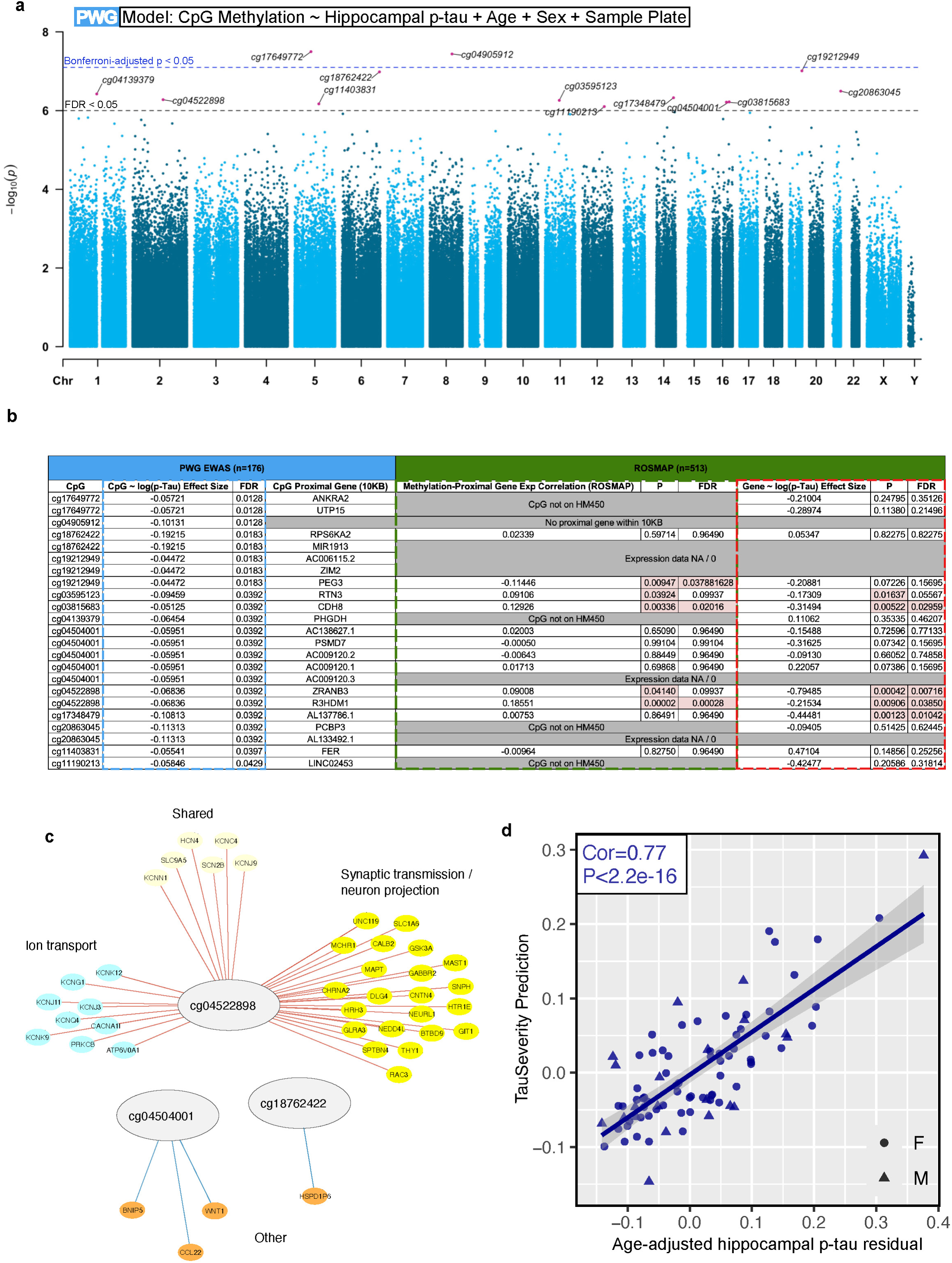
Hippocampal p-tau burden in PART associates with a DNAm signature related to synaptic signaling and cytoskeletal architecture. **a.** Epigenome-wide association study (EWAS) of hippocampal p-tau, covarying for age, sex, and sample plate, reveals 13 novel CpGs (red). Dashed line indicates level of significance after correction for multiple comparisons: False Discovery Rate (FDR) < 0.05 (Black) and Bonferroni-adjusted p-value < 0.05 (Blue). **b.** Table of EWAS CpGs and adjacent genes, along with effect size of hippocampal p-tau burden on CpG methylation (blue outline) in PWG cohort, Pearson correlation coefficients of CpG methylation and gene expression (green outline) in ROSMAP cohort, and effect size of hippocampal p-tau burden on gene expression (red outline) in ROSMAP cohort. Not all CpG:gene pairs could be tested in both cohorts due to lack of detectable RNA expression or absence of a CpG probe on the Human Methylation450 (HM450) beadchip array. n = number of cases. kb = kilobase. **c.** Compared to other EWAS CpGs, methylation at *cg04522898* correlates with the expression of a large network of genes related to synaptic transmission, neuron projection, and ion transport. Red edges = positive correlation, blue edges = negative correlation. **d.** TauSeverity predicts age-adjusted hippocampal p-tau residuals with high accuracy in the PWG cohort. n = 86, Pearson’s r=0.77 and P < 2.2e-16. F = female, M = male.

Given that DNAm often regulates local gene expression programs^39^, we directly evaluated *cis-*regulatory correlations between each of the 13 significant CpGs with gene expression data of all annotated genes within a standard range of 10kB of the CpG probe, using a paired DNAm:gene expression data set from the ROSMAP cohort^35^ (Methods: PART Epigenome Wide Association Study). From the 13 CpGs, we identified 3 significant CpG:gene associations: *cg03815683*:*CDH8*, *cg04522898*:*R3HDM1*, and *cg19212949*:*PEG3* (**Fig. 1b and Extended Data Fig. 2b**). Downregulation of *CDH8* and *R3HDM1*, but not *PEG3*, further correlated with hippocampal p-tau (**Fig. 1b and Extended Data Fig. 2c**). Thus, p-tau-associated methylation at these newly identified loci correlates with the expression of genes that, in turn, are associated with p-tau severity in an independent data set. Methylations of these CpGs were positively correlated with one another (**Extended Data Fig. 3a**), suggesting they may be downstream of a common effector or that they may operate in tandem as part of a concerted epigenetic program.

CpG methylation can also have *trans*-regulatory effects on gene expression^40^ so we expanded our analyses to test CpG:gene associations for all genes, regardless of genomic proximity, with the 13 CpG loci identified in our EWAS (Methods: Differential gene expression analysis). We identified a total of 198 CpG:gene correlations which were enriched in ontology terms related to ion transport, synaptic transmission, and neuron architecture (**Extended Data Fig. 3b**). *cg04522898* correlated with a large network of these genes (**Fig. 1c**), suggesting that methylation at this locus may impact large downstream transcriptional networks related to synaptic biology. Together, these univariate EWAS and CpG:gene findings suggest that p-tau severity in the hippocampus is associated with frontal cortex epigenetic factors in PART, even in the absence of cortical ß-amyloid deposition.

To further investigate the epigenetic correlates of p-tau severity in the PWG cohort, we trained a penalized elastic net (EN) regression model, or “clock”, to predict age-adjusted hippocampal p-tau residuals (Methods). The resultant clock, termed “TauSeverity”, consists of 223 CpGs, including *cg18762422* identified by our EWAS. Our model learned the training data with near certainty and closely predicted hippocampal p-tau residuals in testing data (**Fig. 1d**). As expected, given prior adjustments for age, none of these identified CpGs correlated with chronological age (**Extended Data Fig. 3c**). Notably, in contrast to TauSeverity, existing epigenetic clocks including pan-tissue Horvath^41,42^ or brain-specific Cortical^32^ clocks were not predictive of age-adjusted residual or unadjusted hippocampal p-tau (**Extended Data Fig. 3d**). These results demonstrate that our novel epigenetic clock contains robust frontal cortex DNAm signatures indicative of hippocampal p-tau severity in PART, which are separate from known age-related epigenetic processes^43^.

### Two distinct tau clocks capture the full spectrum of p-tau variation in AD cortex and hippocampus

Unlike PART, AD is also characterized by ß-amyloid inclusions and a more extensive spread of NFTs from beginning in entorhinal cortex and hippocampus before spreading to frontal cortex^8^. Given our evidence of epigenetic contributions to p-tau severity in PART, we next sought to evaluate epigenetic contributions to both p-tau severity and spread across the PART-AD spectrum by analyzing DNAm in the ROSMAP cohort^25^. Due to differences in the DNAm beadchip arrays used in the PWG (Infinium MethylationEPIC) and ROSMAP (Infinium HM450) cohorts, we were unable to directly apply the PWG TauSeverity model to the ROSMAP cohort. Therefore, we trained two new models using the same approach: the ROSMAP “TauSeverity” and “TauSpread” models, related to age-adjusted hippocampal and midfrontal p-tau residuals, respectively. As reported in **Fig. 2a**, we observed accurate predictions of each: TauSeverity (model size: 461 CpGs) and TauSpread (model size: 467 CpGs). Similar to the results in the PWG cohort, the extant cortical and Horvath epigenetic clocks did not accurately predict the p-tau residuals in either region (**Extended Data Fig. 4a**). Neither TauSeverity nor TauSpread models were enriched in aging-associated CpGs within the ROSMAP cohort (**Extended Data Fig. 4b**).

**Fig. 2:**
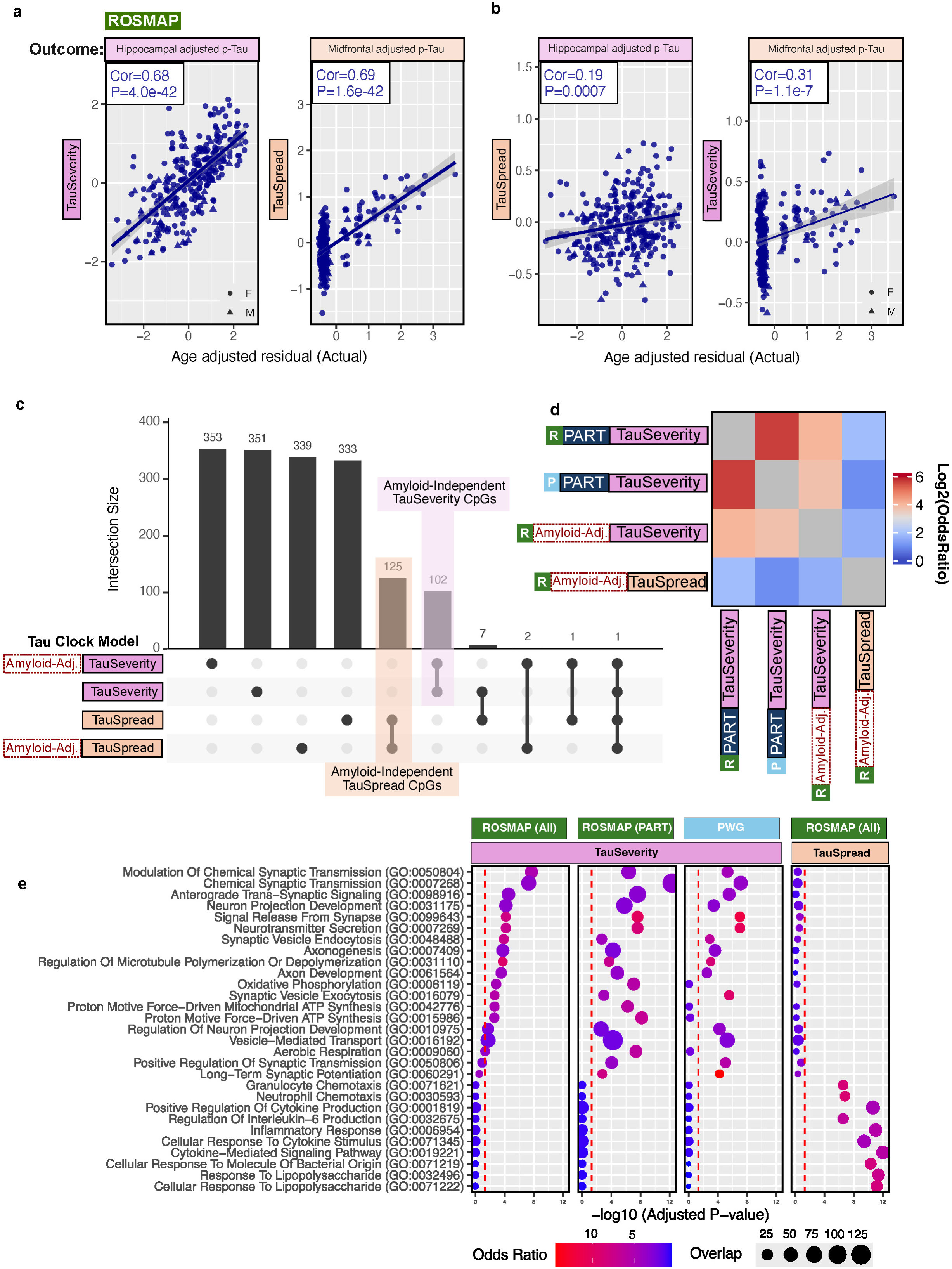
Tau clocks predicts p-tau severity and spread. **a.** TauSeverity predicts hippocampal (left) and TauSpread predicts midfrontal (right) age-adjusted p-tau residuals with high accuracy in the ROSMAP cohort. For TauSeverity, one outlier is not displayed due to axes limits but was included in statistical testing. [hippocampus, TauSeverity] n = 291, Pearson Cor = 0.66, P = 1.342e-37; [midfrontal cortex, TauSpread] n = 291, Pearson Cor = 0.69 and P = 1.68e-42. F = female, M = male. **b.** TauSeverity and TauSpread are region-specific. Models trained to predict hippocampal age-adjusted p-tau residual from TauSpread CpGs (left) do not learn or perform as well as the TauSeverity model shown in Fig. 2A. The same is true for the converse condition (right). [hippocampus, TauSpread] n = 291, Pearson’s Cor = 0.197 and P = 0.0007; [midfrontal, TauSeverity] n = 291, Pearson’s Cor = 0.305 and P = 1.09e-7. F = female, M = male. **c.** Upset plot demonstrating number of disparate and overlapping final feature CpGs in each Tau clock model. The overlapping CpGs from the unadjusted and amyloid-adjusted (Amyloid-Adj.) models are suspected to be amyloid-independent epigenetic modifiers of p-tau in hippocampus (purple) and midfrontal cortex (orange). **d.** Heatmap demonstrates substantial enrichment of methylation-associated gene expression in TauSeverity models developed from PART cases in two independent cohorts. This enrichment is shared with the amyloid-adjusted TauSeverity model from all cases in the ROSMAP cohort, but distinct from the amyloid-adjusted TauSpread model genes from the same set of individuals. R = ROSMAP Cohort. P = PWG group cohort. PART = model trained on PART cases only. Amyloid-Adj. = Genes associated with Amyloid-independent CpG as denoted in Fig 2c. **e.** The feature CpGs of each TauSeverity model are associated with expression of genes enriched for gene ontology (GO) terms related to synaptic transmission, oxidative phosphorylation, and cytoskeletal architecture. In contrast, the feature CpGs of the TauSpread model are associated with expression of genes enriched for GO terms related to inflammation. Statistical testing is detailed in Methods.

We observed only 8 overlapping CpGs between the TauSeverity and TauSpread models, consistent with our hypothesized distinct epigenetic contributions to p-tau severity and spread. Indeed, while both models are moderately correlated (**Extended Data Fig. 4c**), TauSeverity is less accurate at predicting midfrontal p-tau burden, and similarly, TauSpread is less accurate at predicting hippocampal p-tau burden (**Fig. 2b**) highlighting that there are regionally-distinct epigenetic mechanisms that account for variation in p-tau severity and spread.

Whereas 39.4% of the CpGs from the PWG TauSeverity model were missing in the ROSMAP cohort, rendering a cross-validation not possible, only 5.2% and 4.9% of CpGs from the ROSMAP TauSeverity and TauSpread models were missing in the PWG cohort. The ROSMAP TauSeverity model correlated the PWG TauSeverity model (**Extended Data Fig. 5a**) and accurately predicted age-adjusted hippocampal p-tau in the independent PWG cohort (**Extended Data Fig. 5b**). Excluding the missing ∼5% of CpGs from the models did not substantially alter their predictions within the ROSMAP cohort (**Extended Data Fig. 5c**). As expected, the ROSMAP TauSpread model that is predictive of frontal cortex p-tau did not demonstrate an association in the PWG cohort given, by definition, the relatively limited distribution of p-tau in frontal cortex of PART cases.

To validate TauSpread, we turned to frontal cortex DNAm from the publicly-available Mount Sinai Brain Bank data (N=142). Although quantitative histopathology is not available in this dataset, we detected a correlation of TauSpread with Braak NFT stage (**Extended Data Fig. 6**). A similar association was not observed for TauSeverity, as expected given the specificity of the model for hippocampal p-tau only. Taken together, we establish that TauSeverity and TauSpread generalize across cohorts and relate to distinct aspects of overall p-tau burden.

To account for ß-amyloid-dependent processes that may exacerbate p-tau pathology or confound p-tau-specific epigenetic signatures, we re-trained TauSeverity and TauSpread models that, in addition to age, also adjust for total neuritic plaque load (Methods: Tau clock model development). Similar to the respective models that only adjust for age, the ß-amyloid-adjusted models also accurately predict either p-tau severity or spread (**Extended Data Fig. 7a**). Moreover, they have substantial overlap in constituent feature CpGs with models that adjust for age alone (**Fig. 2c** and **Supplemental Table 2**). This suggests that a subset of Tau clock CpGs represent an ß-amyloid-independent epigenetic program that modifies overall p-tau severity and spread.

### TauSeverity and TauSpread epigenetic clocks relates to distinct biological pathways involving synaptic signaling or inflammation

To disentangle the variable epigenetic loci contributing to each of the TauSeverity and TauSpread clock models, we again leveraged integrative paired DNAmgene expression analyses from ROSMAP to identify CpG:Gene pairs for each model (Methods: Differential gene expression analyses). The TauSeverity gene set was largely distinct from the TauSpread gene set, mirroring the regional CpG features selected (**Fig. 2d**, **Extended Data Fig. 7b, and Supplemental Table 3**). Strikingly, despite differences in histopathological and DNAm techniques across PWG and ROSMAP cohorts, a highly overlapping gene profile emerged among TauSeverity models trained separately on PART cases from the PWG cohort, PART cases from the ROSMAP cohort, or all cases from the ROSMAP cohort after adjusting for ß-amyloid. Specifically, the gene ontological profile related to all TauSeverity models was enriched for synaptic transmission, axon biology, and oxidative phosphorylation terms (**Fig. 2e**), suggesting that hippocampal p-tau accumulation in AD and PART appears to be driven by a common process linked with similar epigenetic and transcriptomic features.

In contrast, consistent with spread of p-tau to frontal cortex that occurs primarily in AD, the CpG:Gene correlations for TauSpread were enriched for inflammatory signaling and cytokine production terms. We regressed the expression of the synaptic transmission and inflammation-related genes on regional p-tau and found altered expression of two genes related to synaptic transmission, and five genes related to inflammation after FDR correction (**Extended Data Fig. 7c**). Interestingly, one gene, *kinesin family member 5B* (*KIF5B)*, was common to both midfrontal and hippocampal analyses. Together, these data suggest that hippocampal p-tau severity is associated with altered cortical synaptic activity and connectivity in an ß-amyloid-independent manner both in PART and in AD, while p-tau spread to midfrontal cortex may be precipitated by increased inflammation only in AD.

### Frontal cortex methylations distinguish PART from early AD

Brain resistance, or an individual’s propensity to avoid age-related toxic proteinopathies, and cognitive resilience, or the maintenance of cognitive function despite pathological burden, both contribute to overall cognitive function in aging. PART is often thought of as a model of resistance to AD neuropathologic change, and both low p-tau severity and spread can be thought of as metrics of brain resistance. In view of this, we compared TauSeverity and TauSpread at the extremes of the PART-AD neuropathologic spectrum. Despite significant mean differences between PART and AD for both models, the pathology groups had substantial overlap in each model which limited our ability to discriminate pathology using these two methylation models alone (**Extended Data Fig. 8a**).

Given the significant variability in cognitive function of individuals with comparable p-tau burden^7^, we hypothesized that our neuropathological definitions for PART and AD cases may reflect a combination of brain resistance to pathological accumulation and cognitive resilience to impairments in response to p-tau burden. To investigate this possibility, we developed a support vector machine classifier (the “ResilienceDetector”) to directly stratify Indeterminate cases as “High Resilience” (PART-like) or “Low Resilience” (AD-like) from frontal cortical DNAm (Methods: ResilienceDetector).

We first trained the ResilienceDetector on a random sample of 80% of PART and AD cases from the ROSMAP cohort, and the model predicted PART vs. AD with 74% accuracy (P=.02), and equal sensitivity and specificity, on the remaining 20% of withheld PART and AD cases not seen during feature selection or model training (**Extended Data Fig. 8b,c**). Having validated the approach, we retrained a final model using all PART and AD cases and applied it to cortical DNAm data from 142 individuals defined as Controls or AD in an external cohort maintained by the Mount Sinai Brain Bank (**Extended Data Fig. 8d**). We observed a substantial increase in the percentage of Low Resilience (AD-like) cases with increasing Braak NFT stage (**Extended Data Fig. 8e**), thus validating the classifier’s performance and the reproducibility of its underlying epigenetic AD signature. Most AD cases were classified as Low Resilience (84% AD-like), and most Controls were classified as High Resilience (52% PART-like). Interestingly, a significant subset of AD or Control cases were assigned the opposite resilience category, suggesting potential variability in individual resilience mechanisms despite apparent pathological similarities.

To explore this further, we then applied the ResilienceDetector to stratify “Indeterminate” cases, that fall ambiguously between probable PART and lesser degrees of AD neuropathologic change (CERAD = Sparse, and Braak NFT Stage 0-VI; CERAD = Moderate, and Braak NFT Stage 0-VI; or CERAD = Frequent), from the ROSMAP cohort as High or Low Resilience (**Fig. 3a**). Cases with “No Pathology” (CERAD = “None”, Braak NFT Stage = 0) were excluded due to their limited number (N = 9). High and Low Resilience cases had comparable p-tau or neurtitic plaque burden (**Fig. 3b**) and comparable TauSeverity and TauSpread scores (**Extended Data Fig. 8f**), suggesting that the ResilienceDetector measures an orthogonal epigenetic signature.

**Fig. 3:**
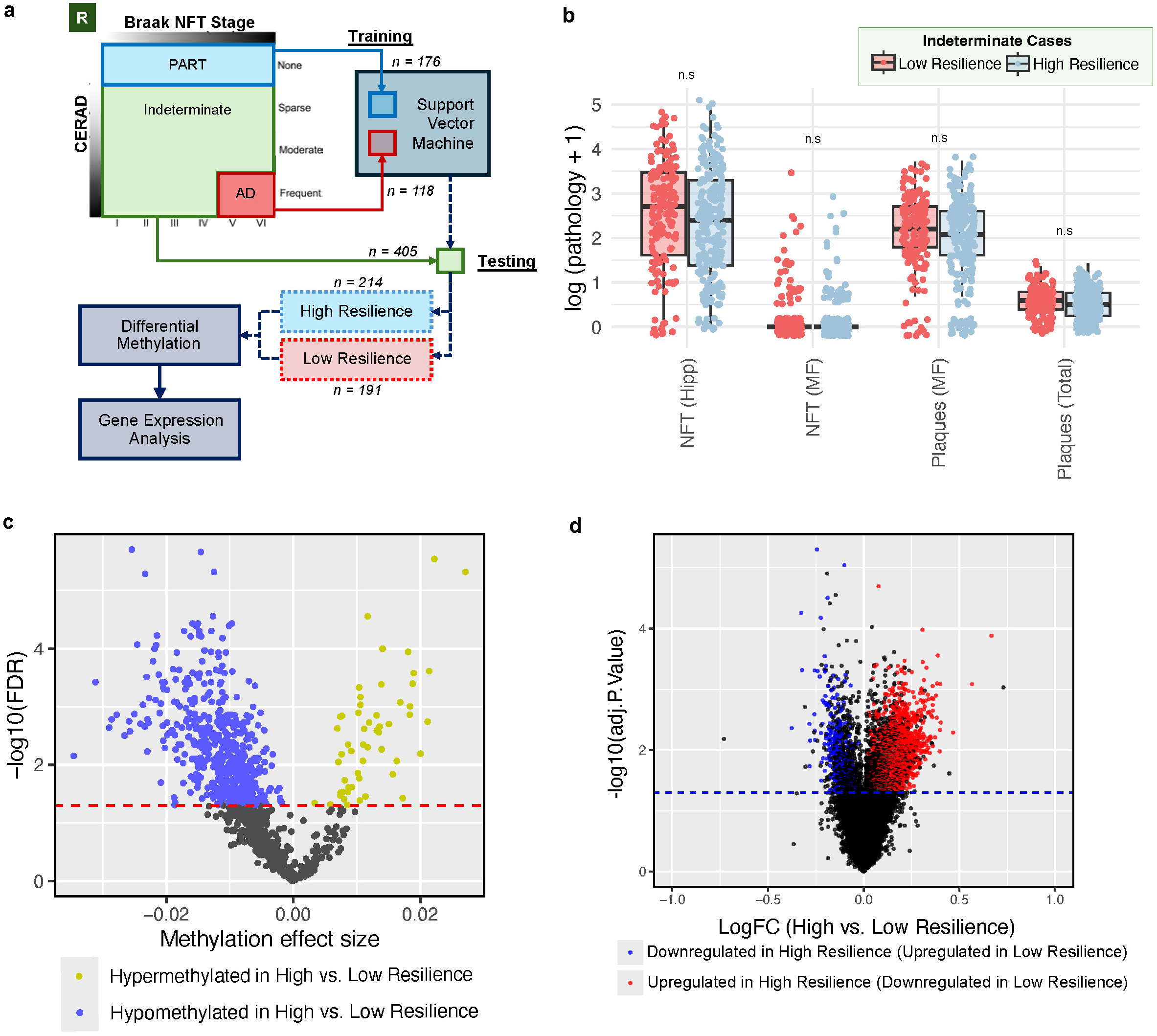
The ResilienecIndicator measures an additional epigenetic signature that distinguishes PART from AD. **a.** Schematic of training and application of the ResilienceDetector. DNAm from cases meeting definition for PART (n=176) or AD (n=118) from the ROSMAP cohort were used to train a support vector machine, which was in turn used to stratify Indeterminate cases (n = 405) from the ROSMAP cohort into High Resilience and Low Resilience cases for subsequent analyses. **b.** Hippocampal (Hipp) and midfrontal (MF) p-tau neurofibrillary tangles (NFTs) and midfrontal and total ß-amyloid neuritic plaques were similar between Low and High Resilience cases. Pairwise t-tests n = 405. n.s = not significant. **c.** Volcano plot of differentially methylated CpGs between High and Low Resilience cases from the ROSMAP cohort (Model: CpG methylation ∼ age + sex + sample_plate + prediction group): n = 405. Highlighted are 570 significant CpGs after correction with an FDR < 0.05 (red line). **d.** Analysis of ROSMAP RNA-sequencing data reveals 4,668 differentially expressed genes between High and Low Resilience cases: n = 405. Highlighted in red (upregulated) and blue (downregulated) are 2,179 of these genes that are also associated with differential-methylation at a feature CpG of the ResilienceDetector and were differentially expressed between confirmed PART and AD cases.

Indeterminate cases that were classified as High or Low Resilience exhibited differential methylation at 570 of the 1,214 total CpGs that are included in the ResilienceDetector model (**Fig. 3c** and **Supplemental Table 4**). Further, we found 4,668 differentially expressed genes between High and Low Resilience cases (Methods: Differential gene expression analyses). Of these, we focused on a subset of 2,179 that were also differentially expressed between confirmed PART and AD as well as correlated with methylation of any of the 570 CpGs that differed between resilience classification groups. (**Fig. 3d** and **Supplemental Table 5**). These methylation-associated High vs. Low Resilience genes had highly concordant effect sizes with those obtained from direct differential gene expression analysis of confirmed PART vs. AD (**Extended Data Fig. 9a**). Genes upregulated in Low Resilience compared to High Resilience cases were mildly enriched in processes of cellular migration and development (**Extended Data Fig. 9b**). Moreover, genes upregulated in High Resilience compared to Low Resilience cases were highly enriched in oxidative phosphorylation and cellular respiration (**Extended Data Fig. 9c**). This may suggest that p-tau associated cognitive impairment may require cellular metabolic dysfunction.

### Methylation models complement in predicting cognitive impairment

P-tau severity^18^ and spread^7^ are both correlated with cognitive impairment. We, therefore, assessed whether methylation signatures measured by TauSeverity, TauSpread, and the ResilienceDetector were related to cognitive impairment in the ROSMAP cohort, measured by mini-mental state examination (MMSE) ante-mortem median of 1.14 (interquartile range: 0.56 - 4.25) years from brain donation. Interestingly, TauSeverity could distinguish individuals with cognitive impairment (MMSE < 24) from those with no cognitive impairment (MMSE ≥ 24, **Fig. 4a**), whereas a prior epigenetic clock could not^31^. On the other hand, TauSpread differentiated individuals with dementia (MMSE < 21), but not from those who have mild (MMSE 21-23) or no cognitive impairment. Together, the two Tau clock models can separate individuals along the spectrum of cognitive impairment, likely with TauSeverity more sensitive to impairment in any cognitive domain and TauSpread more sensitive to the number of cognitive domains involved.

**Fig. 4:**
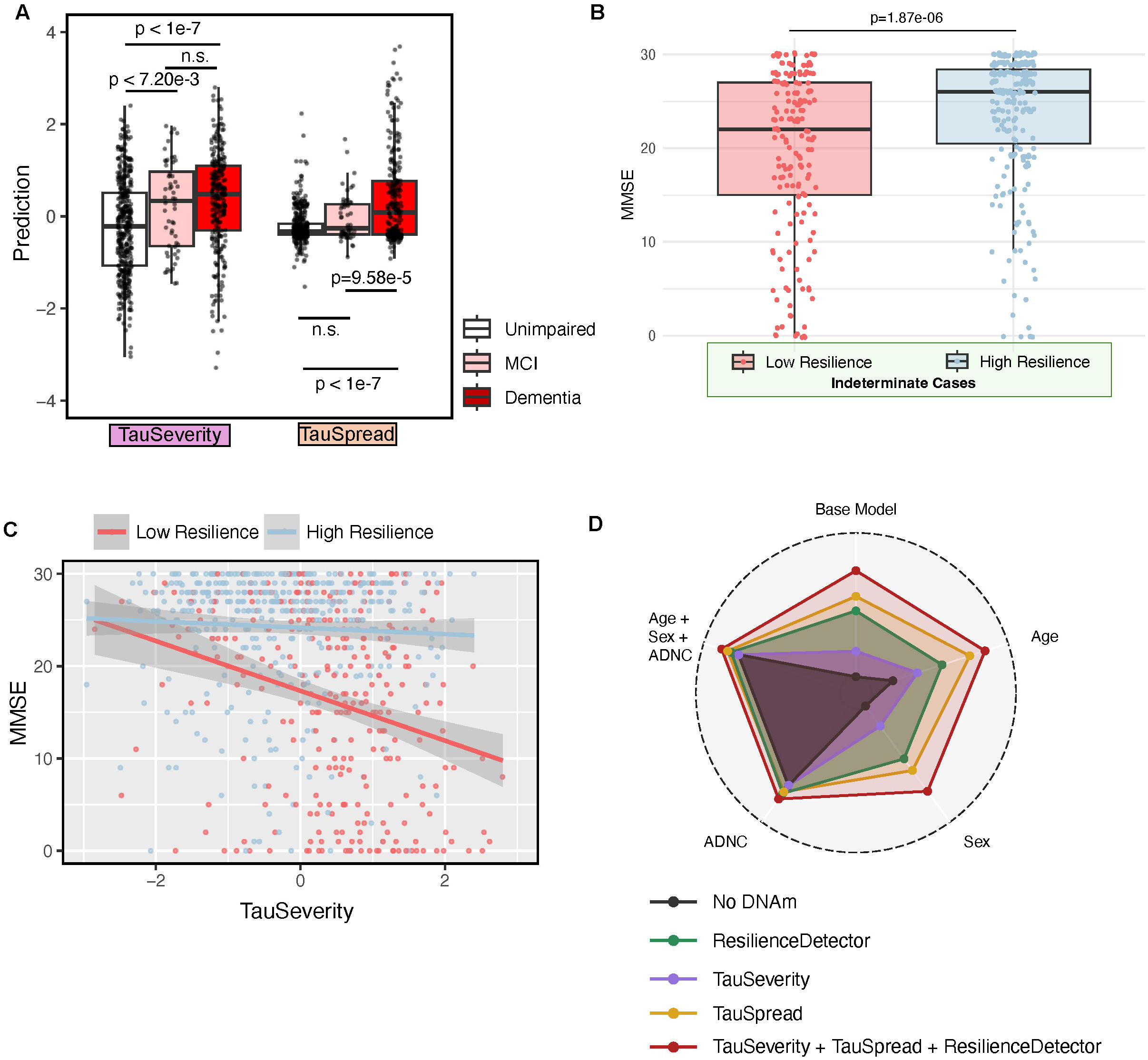
TauSeverity, TauSpread, and the ResilienceDetector estimate cognitive impairment better than age or neuropathologic change alone. **a.** Increasing TauSeverity (left) and TauSpread (right) in the ROSMAP cohort are associated with decreasing performance on the mini-mental status examination (MMSE) and, therefore, increasing cognitive impairment. For TauSeverity, one outlier is not displayed due to axes limits but was included in statistical testing. Statistical test was a one-way ANOVA followed by Tukey’s HSD test: n = 707. **b.** Of the Indeterminate cases classified by the ResilienceDetector, Low Resilience cases exhibit lower MMSE score and, therefore, greater cognitive impairment compared to High Resilience cases in the ROSMAP cohort. Statistical test was a two-tailed *t*-test: n = 405. **c.** There is an inverse correlation of TauSeverity and MMSE score with a larger slope of the effect in Low compared to High Resilience cases. For TauSeverity, one outlier is not displayed due to axes limits but was included in statistical testing. [High Resilience] n= 418, Cor = −0.055, and P = 0.258; [Low Resilience] n= 288, Cor = −0.28, and P = 1.59e-06. **d.** Radar plot depicts the variance explained in cognitive impairment by by different models. The Base Models include: No DNAm, TauSeverity alone, TauSpread alone, the ResilienceDetector alone, and a full DNAm Base Model combining all three (TauSeverity + TauSpread + ReselienceIndicator). Each of these models is also evaluated with additional covariates: Age, Sex, and AD Neuropathologic Change (ADNC). Notably, the variance explained by Age + Sex + ADNC is nearly equivalent to that explained by the full DNAm Base Model, suggesting that these epigenetic predictors capture biological variance comparable to traditional demographic and neuropathologic measures.

Despite similarities in p-tau burden in prediction groups identified by the ResilienceDetector, High Resilience individuals scored higher on the MMSE than Low Resilience individuals (**Fig. 4b**). Interestingly, the effect of TauSeverity on MMSE is more pronounced in the Low Resilience individuals than in High Resilience individuals (**Fig. 4c**). Therefore, while PART and AD share similar hippocampal p-tau, susceptibility to metabolic dysfunction and immune cell infiltration may result in more dramatic cognitive impairment in AD. As a corollary, decreased metabolic dysfunction in PART and lack of inflammation may mediate more resilient cognitive aging. Finally, to quantify the relative contributions of each model in predicting cognitive resilience, we constructed several multiple regression models and evaluated the variance explained in MMSE (**Fig. 4d**). Age alone explained only 6.4% of the overall variance in MMSE performance. Among single methylation models, TauSpread performed best (22.4%), followed the the ResilienceDetector (18.3%), and TauSeverity (7.1%). When combined, the three DNAm predictors explained 29.5% of the variance, greater than any model alone, suggesting they capture distinct and largely non-overlapping biological pathways influencing cognitive decline in aging and AD.

Strikingly, the variance explained by the DNAm-only model (TauSeverity + TauSpread + ResilienceDetector) was nearly equivalent to that of a traditional model that included age, sex, and AD neuropathologic change (ADNC; 29.8%). Combining the traditional and DNAm models only modestly improved performance (34.8%), the marginal gain reinforces that the bulk of predictive power resides in the methylation models themselves and underscores their potential as robust molecular indicators of pathology and resilience in aging and AD.

## DISCUSSION

This study establishes an epigenetic landscape of PART. By carefully disentangling correlations between p-tau, ß-amyloid, and DNAm, across a neuropathological spectrum of PART and AD we define two epigenetic clocks linked with regional p-tau pathology. Two key processes – synaptic signaling and neuroinflammation – differentially associate with early hippocampal p-tau severity and its later spread to the frontal cortex, respectively. Critically, these findings highlight important molecular insights into age- and AD-associated p-tau biology, discussed below in detail.

### TauSeverity and TauSpread are novel methylation-based measures that dissociate two axes of p-tau burden

TauSeverity and TauSpread models delineate two epigenetic axes related to p-tau severity and spread in PART and AD, and outperform existing epigenetic clocks in accurately predicting age-adjusted p-tau. These clocks have several key properties that distinguish them from more non-specific measures of epigenetic aging. First, they reflect brain-specific biological processes that are age-independent. Second, TauSeverity focuses on within-region severity in p-tau burden whereas prior studies focused on DNAm associations with Braak NFT stage, a measure of the anatomic extent of p-tau spread. Third, our approach results in distinct clocks reflecting unique aspects of p-tau biology, whereas most epigenetic models aggregate different tissues and pathologies, attributing them to a single age-related biological process. Critically, this is the first demonstration that subsets of cortical DNAm loci can differ with regards to local and remote pathology, and that both can be measured simultaneously.

A key difference between PART and AD is the relative sparing of p-tau inclusions in the frontal cortex in PART. TauSpread and the ResilienceDetector could distinguish individuals with dementia from those with mild cognitive impairment, likely leveraging an AD-specific epigenetic signature related to a T-cell and macrophage-mediated axis of inflammation. Our findings from these models add to the growing evidence that PART and AD may have distinct molecular bases. Based on the current observations and our prior genetic studies^14,23^, we posit that PART may reflect both a model of resistance to p-tau spread and a model of resilience despite p-tau severity that are moderated through both epigenetic and genetic factors.

### DNAm can be associated with hippocampal p-tau pathology in aging independently of ß-amyloid

In conducting the first epigenome wide association study of PART, we identified 13 novel CpGs and nominated three candidate genes (*CDH8*, *R3HDM1*, and *PEG3*) as potential *cis* mediators of tau-associated DNAm. These genes have been linked to altered neuronal connectivity^44–47^. For instance, the *R3HDM1* gene has been linked with age with DNAm at *cg04522898* in whole blood.^48^ The *R3HDM1* locus harbors the *pri-miR-128-1* gene^49^, and miR-128 has been implicated in synaptic plasticity and cellular survival by additional epigenetic mechanisms^50–52^. Indeed, when we expanded our analyses to assess additional CpG-gene expression relationships, we found 198 genes that were differentially expressed in relation to differential methylation of at least one of the 13 CpG sites. A majority of these transgene expression effects, including on *MAPT*, were related to methylation at *cg04522898* which lies within the *R3HDM1* locus and may relate to miR-128 function. While functional validation is required to directly investigate the potential mechanistic role of these genes in tau biology, this study points to several novel genomic loci that may contribute mechanistically to p-tau severity in hippocampus.

### Synaptic signaling is a shared determinant of hippocampal p-tau severity in both PART and AD

Synapse dysfunction is a key early hallmark of AD^53,54^. Similarly, synaptic dysregulation may occur in aging and in PART, though there is less consensus on specific molecular etiologies^55–59^. In this study, we demonstrate strong associations between p-tau pathology and synaptic transmission genes. Interestingly, we also find that increased expression of *kinesin family member 5B* (*KIF5B)*, a microtubule motor associated with lysosomal function and mitochondrial localization, positively correlates with both p-tau severity and spread^60,61^. This observation converges with a similar observation in the P301S murine model of tauopathy^62^. Therefore, abnormal cytoskeletal trafficking may synergistically connect to abnormal tau pathology.

Our findings independently corroborate but are unique from prior transcriptomic studies in AD brain, that similarly implicate synaptic dysregulation in AD^63–66^. We first identify DNAm differences associated with hippocampal p-tau and then relate differential methylation to differential gene expression. Moreover, in our study, we focused on the subset of CpGs and associated genes that are likely to be ß-amyloid-independent p-tau modifiers. Lastly, ours is the only study to specifically analyze individuals with PART, and we find that differential methylation converged on overlapping synapse-related genes in both the PWG and ROSMAP cohorts.

### PART represents a cognitively resilient form of aging

Our machine learning-based analyses of DNAm suggest that AD and PART both involve dysregulation of synaptic transmission and plasticity, raising the possibility that PART can set the stage for the development of AD. Individuals with lower p-tau burden than expected for their age exhibit an epigenetic signature indicative of preserved synaptic function, which may confer protection against the onset of ß-amyloid aggregation. In other words, we suspect that a component of the age-associated AD risk is due to underlying PART. Ours is the largest and only evaluation of DNAm in a well-characterized cohort of individuals with PART, and further research is required to determine whether ß-amyloid-independent brain DNA methylation differences consistently predict susceptibility to hippocampal p-tau severity in additional datasets.

Importantly, our ResilienceDetector uses an epigenetic signature distinct from the Tau clock models to stratify cases with indeterminate pathology. Gene ontology analysis identified a role for increased metabolic capacity in High Resilience cases and increased cell migration and transcription in Low Resilience cases. Given numerous other reports of inflammation-associated transcriptomic changes in brain^67–71^, the latter may represent an effect of infiltrating inflammatory cells, and this can be evaluated in more detailed studies using single-cell RNA sequencing or spatial transcriptomics.

Both p-tau and neuritic plaque burden were similar in High and Low Resilience cases. Strikingly, High Resilience cases exhibited better cognitive performance. Within High Resilience cases, there was still an appreciable decline in cognitive performance associated with higher TauSeverity; however, this effect was far more pronounced in Low Resilience cases. This suggests a potential additive effect of synaptic dysregulation and inflammation on cognition in AD.

Importantly, our three novel methylation models relate to distinct underlying biology and synergistically measure cognitive impairment, as evidenced by the increased variance explained in MMSE performance with a summative model. TauSeverity can distinguish the cognitively impaired from unimpaired, and it appears to model synaptic pathophysiology shared by PART and AD. Together, these findings lend credence to the hypothesis that age-associated cognitive impairment due to hippocampal synaptic dysfunction may precede and be requisite for typical AD.

From here, despite similarities in pathological burden, individuals with early AD begin to diverge from PART in a manner detectable by our ResilienceDetector and related to an epigenetic signature of brain metabolic failure. Finally, as p-tau disseminates widely through the neocortex, TauSpread increases, distinguishing MCI from dementia-level of impairment and relating to T-cell-mediated inflammation. This absence of an inflammatory signature in PART cases is potentially neuroprotective and indicates a new pathophysiological axis differentiating PART from AD.

### Caveats to Consider and Overall Conclusions

Critically, while unable to directly cross-validate EWAS CpGs across PWG and ROSMAP cohorts, we were able to validate our ROSMAP TauSeverity, TauSpread, and ResilienceDetector epigenetic models in multiple cohorts. Moreover, there was substantial intersection of the differentially expressed genes regulated by the respective feature CpGs of TauSeverity models separately trained in the PWG and ROSMAP cohorts despite the notable methodological differences between the cohorts.

First, the two studies ascertain DNAm using two similar but distinct array chips, which include nonoverlapping sets of CpG sites. We were able to evaluate the ROSMAP model in the PWG cohort, but not the converse. Second, the two studies used different immunohistochemistry methods that vary in their p-tau affinity. Specifically, in ROSMAP, NFTs were identified by silver stain, which is more sensitive to mature NFTs, whereas in PWG, NFTs were identified by AT8 antibody, which detects both mature and immature p-tau assemblies. Despite these technical differences at the levels of microarray and histopathology, the convergence of the models at the gene expression level suggests that they are two convergent views of the same underlying biology.

Despite these caveats, our epigenetic models clarify molecular similarities and differences between AD and PART and highlight a role for age-independent interindividual variation in hippocampal p-tau severity as a separate risk factor for AD and age-associated cognitive changes more broadly.

### INDEX OF SUPPLEMENTAL MATERIALS TOO LARGE TO UPLOAD AS PDF

**Supplemental Table 2: Tau clock feature CpGs.**

**Supplemental Table 3: Tau clock-associated differentially-expressed genes.**

**Supplemental Table 4: ResilienceDetector feature CpGs.**

**Supplemental Table 5: ResilienceDetector-associated differentially expressed genes.**

## METHODS

### Patient Selection and Cohorts

Fresh-frozen brain tissue was obtained from the contributing centers in the PART Working Group (PWG) as previously described^23^. Inclusion criteria were individuals with normal cognition, mild cognitive impairment (any type) and dementia. Cognitive status was determined either premortem or postmortem by a clinical chart review, mini-mental score, or clinical dementia rating^72,73^. Neuropathological assessments were performed at the respective centers using standardized criteria including Consortium to Establish a Registry for Alzheimer’s Disease (CERAD) neuritic plaque assessment and Braak neurofibrillary tangle staging.^8,74^ In addition, formalin fixed paraffin-embedded tissue sections were obtained and reevaluated by the study investigators to confirm the lack of Aβ and degree of PART p-tau pathology as previously described^24^. Clinical exclusion criteria were motor neuron disease, parkinsonism, and frontotemporal dementia. Neuropathological exclusion criteria were other degenerative diseases associated with NFTs (i.e., AD, progressive supranuclear palsy [PSP], corticobasal degeneration [CBD], chronic traumatic encephalopathy [CTE], frontotemporal lobar degeneration-tau [FTLD-tau], Pick disease (PiD), Guam amyotrophic lateral sclerosis/parkinsonism–dementia, subacute sclerosing panencephalitis, globular glial tauopathy). Individuals with aging-related tau astrogliopathy (ARTAG) were not excluded^75^. Characteristics and procedures of the Religious Orders Study and Memory and Aging Project (ROSMAP) were as previously described^76^. Characteristics and procedures of the Mount Sinai Brain Bank (MSBB) cohort were as previously described^77^.

### Case definitions and histopathology

To allow for comparisons between cohorts and to minimize confounding due to overlapping pathologies, we limited our analyses using the following case definitions, using variables common across PWG and ROSMAP cohorts. We conserve our definition of PART to “definite PART” defined as Braak NFT stage I-IV and CERAD = None^6^. For AD, we restrict to cases that meet Braak NFT stage VI-V and CERAD = Frequent^20^. Remaining cases were termed Indeterminate. Digital histopathology in the PWG cohort was performed as previously described^78^. Briefly, stains were performed on 4 µm-thick formalin-fixed paraffin-embedded (FFPE) sections stained with AT8 antibody. Sections from the body of the hippocampus were targeted, but this neuroanatomical landmark was not represented in all sections, and there was some variability noted with regard to representation along the anterior–posterior axis. Whole slice images were scanned using an Aperio CS2 (Leica Biosystems, Wetzlar Germany) digital slide scanner at 20 × magnification, and neurofibrillary tangle density was calculated via a SegNet model architecture as previously detailed^34^. Manual histopathology assessment in the ROSMAP cohort was performed by independent neuropathological assessment of silver-stained tissue, as previously described^79^. To validate the PART-AD classifier, we tested it on cases from the MSBB cohort, which the investigators previously defined as Control (Braak NFT Stage 0-II), or AD (Braak NFT Stage III-VI).

### DNAm data generation

In the PWG cohort, DNA was extracted from the frontal cortex. Bisulfite converted DNA was profiled using the Illumina Infinium MethylationEPIC bead chip array at the Center for Applied Genomics core at the Children’s Hospital of Philadelphia. In the ROSMAP cohort, DNA extraction and methylation profiling using the Illumina Infinium Human Methylation450 bead chip assay was performed as previously described^25^. Raw data from both cohorts was used and subjected to the same standard manufacture recommended preprocessing and quality control pipeline using the SeSAMe R package (version 1.22.0). Beta values were extracted using the openSesame function with default parameters as previously described^80^.

### Unsupervised clustering analysis

CpGs with >50% missingness across all samples were removed and remaining missing values were imputed using the beta value mean from non-missing samples. tSNE analysis was performed using the Rtsne package (version 0.16) with a perplexity of 30.

### Cell type deconvolution

Beta matrices for PWG and ROSMAP cohorts were filtered for common CpGs with coverage across 75% or more of samples in both cohorts. A reference matrix for 20 major brain cell types was constructed by performing one vs. all non-parametric analyses of pseudobulk methylomes obtained from publicly availably single cell WGBS data^36–38^. Reference-based cellular deconvolution was performed using the EpiDISH R package (version 2.16.0) with the robust partial correlations (RPC) method. Principal component analysis was performed with the prcomp function from the stats package (version 4.4.0) using the cell type proportions as input features. Box and whisker plots depict median, interquartile range and with outliers indicated if greater than 1.5x interquartile range.

### PART Epigenome Wide Association Study (EWAS)

CpG methylation (CpGm) was regressed on logarithmically-transformed hippocampal p-tau (hip_p-tau) with age, sex and sample plate added as covariates. [Model: CpGm ∼ age + sex + sample plate + *log*(hip_p-tau + 1)] Modelling was performed using the DML() function from the SeSAMe R package (version 1.22.0). P values for each modelled CpG were corrected for multiple comparisons by the false discovery rate (FDR) method, and only those with FDR < .05 were considered for further analysis. Gene annotations were retrieved using the sesameData_txnToGeneGRanges function from the sesameData package (version 1.21.9) and intersected with EWAS hit CpGs (expanded by 10kb) using the subsetByOverlaps function from the GenomicRanges package (version 1.57.1).

### Tau clock model development

Each tau clock is a novel statistical model that uses methylation at a distinct set of feature CpGs to predict an age-adjusted regional p-tau residual. To calculate the age-adjusted p-tau residual, logarithmically-transformed hippocampal or midfrontal p-tau was regressed on age [*log*(p-tau + 1) ∼ age].

Amyloid-adjusted tau clocks similarly predicts an age-and amyloid-adjusted p-tau residual. To calculate age- and amyloid-adjusted residuals, logarithmically-transformed total brain neuritic plaques were added as a covariate [*log*(p-tau + 1) ∼ age + *log*(plaque + 1)].

For each model, training data were balanced to include equal numbers of male and female samples (80% of lesser-represented sex), and remaining samples were used for testing. The target variable was age-residualized hippocampal or midfrontal p-tau for TauSeverity and TauSpread, respectively. For any given cohort and target variable, an elastic net (EN) regression of p-tau residuals on input CpG feature methylation beta values was performed using the cv.glmnet function from the glmnet package (version 4.1.8) with an alpha parameter of 0.5 and 10-fold cross validation.

For each target, CpG features were rank ordered according to the unadjusted P-value from running a univariate EWAS [Model: CpGm ∼ age + sex + sample plate + *log*(variable + 1)] on training data only. Feature sizes were scanned from the top 2,000-to top 30,000-ranked CpGs, and models with the most highly correlated predictions to actual values on testing data were selected for further analysis.

For TauSeverity, one outlier in the ROSMAP cohort was 6 standard deviations below the cohort mean. This outlier was excluded from visualization but was included in all statistical analyses. To confirm the specificity of each tau clock model in the ROSMAP cohort, separate models were trained to predict one region’s p-tau residual using the opposite models’s feature CpGs.

Having internally validated the approach, final models for TauSeverity and TauSpread were trained after performing feature selection with all cases from the ROSMAP cohort and applied to DNAm from cases in the PWG and MSBB cohorts, excluding CpGs not measured in the PWG cohort as described in the main text.

### Epigenetic age analysis

Missing values from processed CpG beta matrices were imputed using the row mean from non-missing samples, and epigenetic age estimates (mAge) were computed using the methyAge function from the dnaMethyAge package (version 0.2.0)^81^. Age acceleration was computed by calculating the residual of epigenetic age regressed onto the chronological age [mAge ∼ Age]. Pearson correlations were computed between the age acceleration residuals and p-tau load or p-tau residual.

### ResilienceDetector development

Initial models were trained on 80% of randomly sampled AD cases (n=95) and 80% of randomly sampled PART cases (n=141). For feature selection, CpG methylation was regressed on pathology group (PART vs. AD) with age, sex and sample plate added as covariates (Model: CpGm ∼ age + sex + sample plate + pathology) and the top 1000 CpGs associating with pathology group by P value were selected for model features. PART cases were those with Braak staging I-IV and a CERAD score of None, while AD cases were those with Braak stage V-VI, CERAD score of Frequent. Model performance was evaluated in the held out 20% of cases from each pathology group. For the final model, all PART and AD cases were used with the same feature selection process. Cases were predicted as either PART-like (High Reslience) or AD-like (Low Resilience). Feature selection modelling was performed using the DML function from the SeSAMe R package (version 1.22.0) and support vector machine classifiers were trained using the svm function (kernel=”linear”) from the e1071 package (version 1.7-14). For CpGs with missing values during model training or testing, the mean over all other samples was used for imputation. Performance statistics were computed using the Caret library (version 6.0-94)^82^.

### Differential gene expression analysis

Fragments per kilobase of transcript per million mapped reads (FPKM)-normalized gene expression data from the ROSMAP cohort was log transformed and tested for correlation with methylation of PART EWAS CpGs, TauAge feature CpGs, or PART-AD Classifier feature CpGs. Correlations were FDR-adjusted and CpG methylation:gene expression pairs with an absolute effect size value >= .2 and FDR < .0001 were filtered for further analysis. Only cases where both DNAm and RNA-sequencing data were available were included. To identify genes differentially expressed between AD:PART cases or High:Low Resilience cases, linear models were fit for each gene using the lmFit function from the Limma R package (3.58.1). The variance of a gene was stabilized using the eBayes function. DEGs were identified with a moderated t-test using the decideTests function with default parameters. To identify genes associated with p-tau pathology, gene expression for each gene was regressed on log transformed p-tau using the lm() function and genes with an FDR<.05 were considered for further analyses. All gene ontology analyses were performed using Enrichr^83–85^.

### Statistical testing (High vs. Low Resilience)

Standard two-tailed t-tests were performed using the t.test function (stats 4.4.0) between High and Low Resilience cases to compare mean TauSeverity, TauSpread, NFTs (hippocampal and midfrontal), neuritic plaques (midfrontal, total) and MMSE with corrections for multiple comparisons using the FDR method. Box and whisker plots depict median, interquartile range and with outliers indicated if greater than 1.5x interquartile range.

### Multiple Regression model

To model cognitive impairment, linear regression models were developed using patient covariate and epigenetic prediction model scores as predictors. MMSE scores were regressed on each of the variables shown in Fig. 4D (No DNAm, TauSeverity, TauSpread, ResilienceDetector, or all methylation models) either without additional covariates (Base Model) or with a combination of Age, Sex, or AD Neuropathologic Change (ADNC) using the lm() function in R version 4.4.0. The adjusted R squared value was extracted for each regression model and plotted using the ggradar package (version 0.2)

## Data Availability

All data produced in the present study are available upon reasonable request to the authors.

## Acknowledgements

We wish to thank all the individual study volunteers, tissue donors, and caregivers with whom this work would not be possible. This work was supported by funding from the National Institutes of Health (R01 AG054008, R01 AG060961, R01 AG062348, R01AG066152, P30AG072979, R01 NS095252, R01 NS086736, R35GM146978, K01 AG070326, P01AG066597, P01AG084497, P30 AG066514, T32AG076411, UE5NS065745), CurePSP 685-2023-06-Pathway, the Rainwater Charitable Foundation / Tau Consortium, Karen Strauss Cook Research Scholar Award, and Penn Institute on Aging. The results published here are in whole or in part based on data obtained from the AD Knowledge Portal. Study data were provided by the Rush Alzheimer’s Disease Center, Rush University Medical Center, Chicago. Data collection was supported through funding by P30AG10161, P30AG72975, R01AG15819, R01AG17917, R01AG30146, R01AG36836, U01AG46152, U01AG61356, and the Illinois Department of Public Health (ROSMAP). Additional phenotypic data can be requested at www.radc.rush.edu. Brain banking and neuropathology assessments for the MSBB cohort were supported by NIH grants AG02219, AG05138, and MH064673 and the Department of Veterans Affairs VISN3 MIRECC.

## Resource Availability

*<u>Lead Contact: Corey T. McMillan, PhD</u>*

*<u>Data and Code Availability:</u>*

The data presented from the multi-institutional PART working group are available on reasonable request from the corresponding author. The data are not publicly available due to privacy or ethical restrictions. The data presented from ROSMAP were provided by the Rush Alzheimer’s Disease Center, Rush University Medical Center, Chicago, via the AD Knowledge Portal and are available on reasonable request www.radc.rush.edu. The data presented from the Mount Sinai Brain Bank cohort are publicly available. Code used to generate the TauAge and PART-AD Classifier models as detailed in the methods will be maintained on GitHub: https://github.com/pennbindlab

## Ethics Statement

The data presented from the multi-institutional PART working group are from post-mortem tissue donated to the respective institution by the individual’s next of kin. All material is de-identified. Research with de-identified autopsy material does not meet the federal regulatory definition of human subject research as defined in 45 CFR part 46 has been deemed exempt from review by the Institutional Review Board of Mount Sinai School of Medicine. The data presented from the Religious Orders Study and Memory and Aging Project are available by controlled access, and use in this study was reviewed by the Resource Sharing Committee of the Rush Alzheimer’s Disease Center. The data presented from the Mount Sinai Brain Bank cohort are publicly available. I confirm that all necessary patient/participant consent has been obtained and the appropriate institutional forms have been archived, and that any patient/participant/sample identifiers included were not known to anyone (e.g., hospital staff, patients or participants themselves) outside the research group so cannot be used to identify individuals.

## Author Contributions

D.C.G. and A.R.W. contributed equally to this study. C.T.M. and W.Z. contributed equally to this study. Conceptualization: C.T.M. and W.Z. Study design and methodology: D.C.G., A.R.W., C.T.M., and W.Z. Collection of data: J.F.C., K.F., D.A.B., and E.B.L. Data Analysis: D.C.G., A.R.W., and N.D. Manuscript writing: A.R.W., D.C.G., C.T.M., and W.Z. Manuscript editing: A.R.W., D.C.G., N.D., L.P.S., E.B.L., D.A.W., D.A.B., C.T.M., and W.Z. Resources and funding: C.T.M. and W.Z.

---

**Extended Data Fig. 1:**
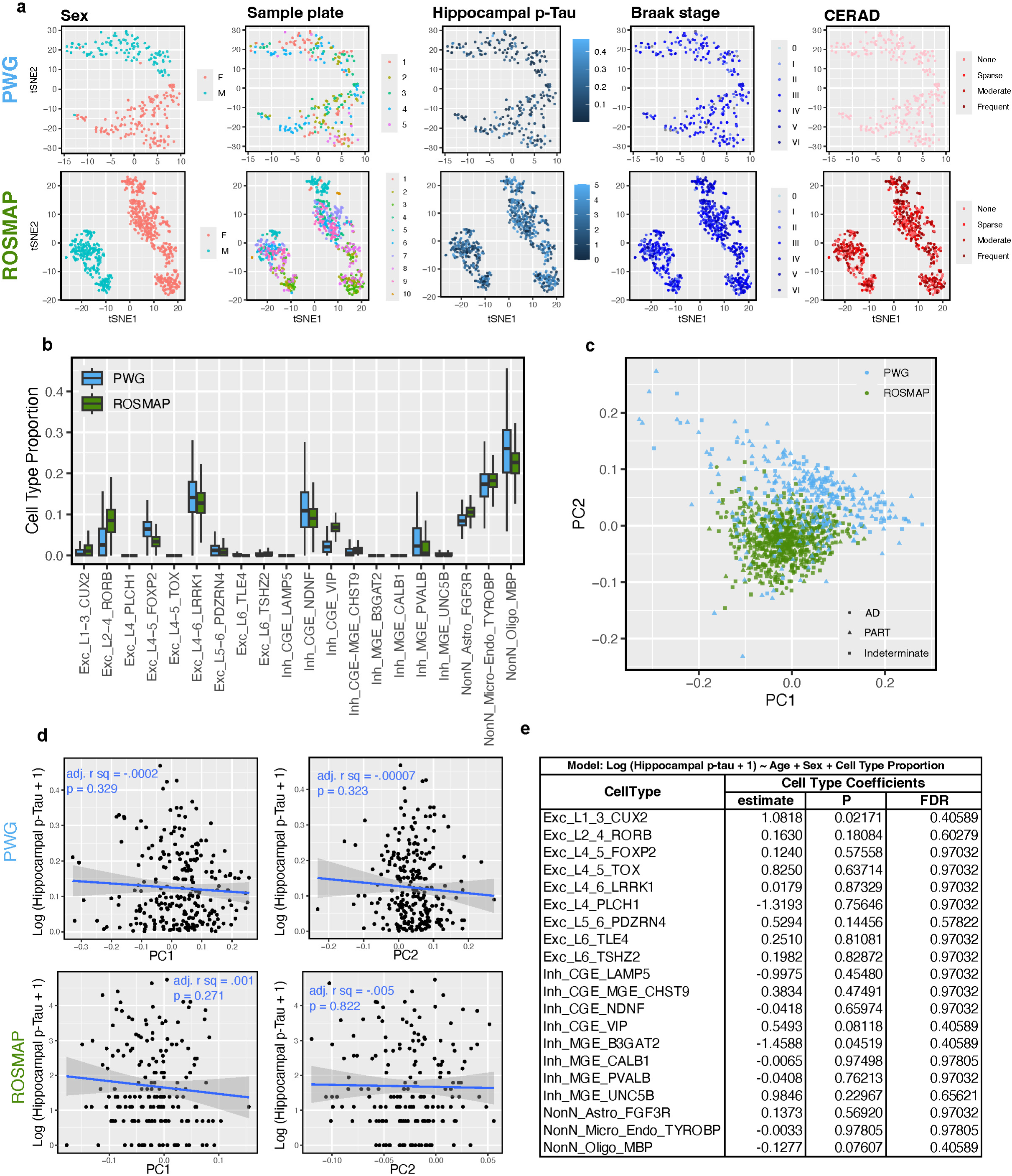
Methylation structure. **a.** T-distributed stochastic neighbor embedding (t-SNE) clustering of DNAm in both cohorts shows organization of data by sex and sample plate, but not hippocampal p-tau severity, Braak NFT stage, or CERAD score. **b.** Cell type proportion estimates for 20 major brain cell types from methylation-based deconvolution in PWG and ROSMAP. **c.** Principal component analysis using estimated cell type proportions demonstrates mild differences in cell type composition between two cohorts. **d.** Regressing hippocampal p-tau on cell type proportion principal components demonstrates that p-tau is not significantly associated with an individual sample’s cell type composition in either cohort. **e.** Regressing hippocampal p-tau on cell type proportions, covarying for age and sex, did not reveal an association for any cell type in PWG cases. FDR = False Discovery Rate

**Extended Data Fig. 2:**
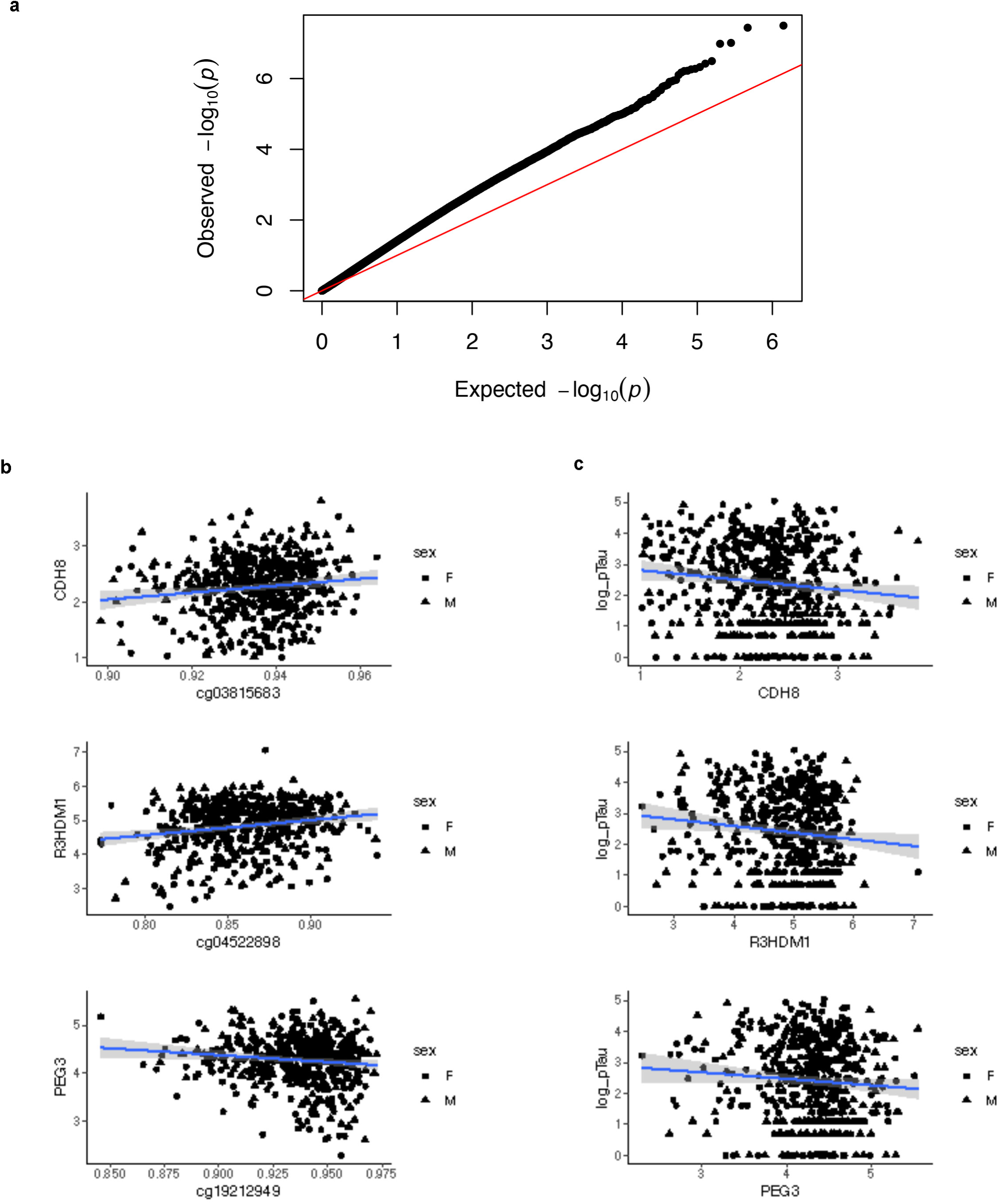
Identification of PART EWAS hits and cis-regulated genes. **a.** Quantile-Quantile (QQ) plot of CpG probes from EWAS in Fig. 1A. Unity line is displayed in red. **b.** Correlation of EWAS hits and expression of candidate cis-regulated genes. Statistics are shown in Fig. 1B. F = Female, M = Male. **c.** Correlation of CpG-associated gene and hippocampal p-tau. Statistics are shown in Fig. 1B. F = Female, M = Male.

**Extended Data Fig. 3:**
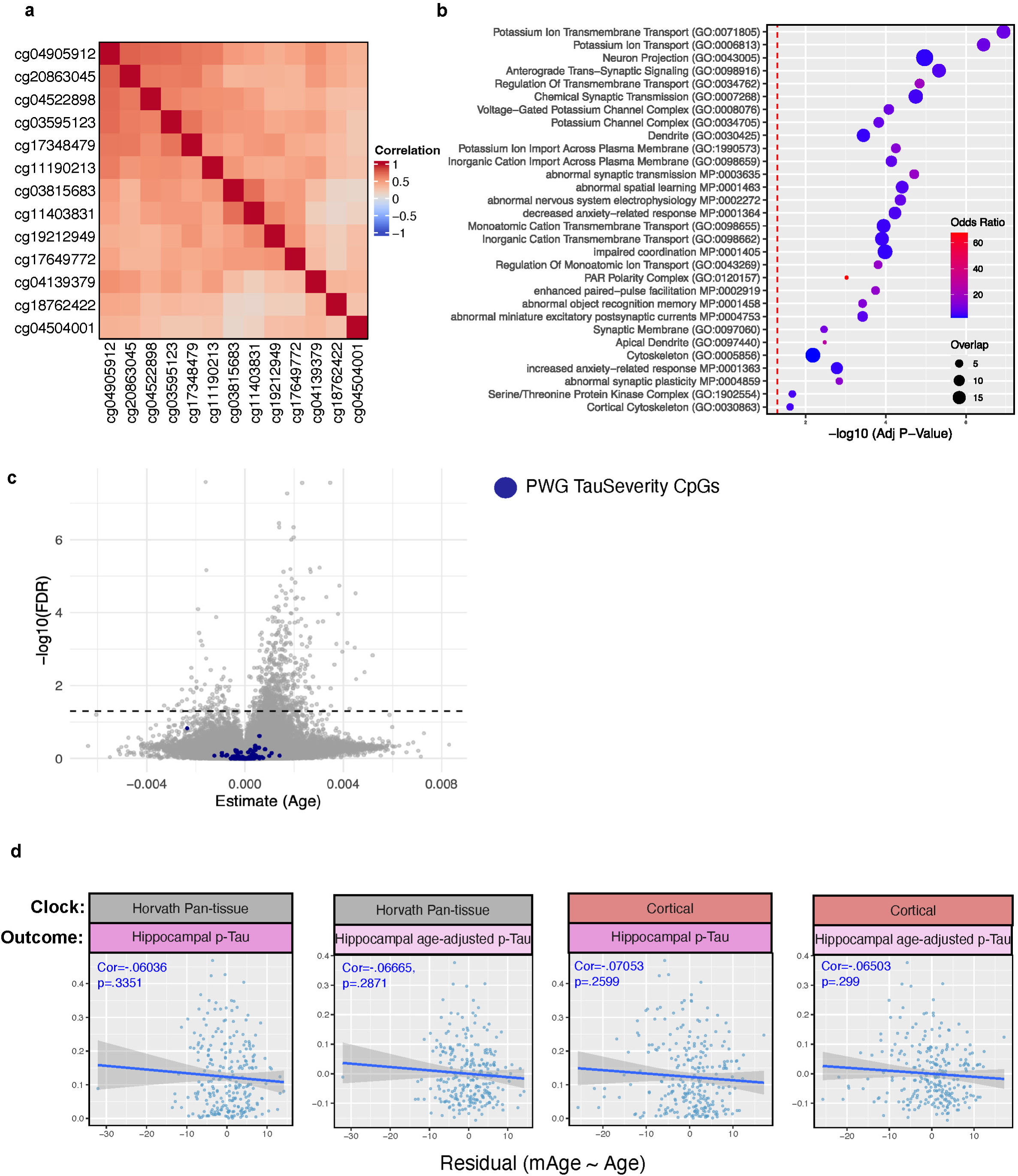
Characterization of PART EWAS hits and cis-regulated genes. **a.** Heatmap demonstrates that PWG EWAS CpGs are correlated with one another (Pearson’s correlation). **b.** EWAS CpG methylations are correlated with expression of genes enriched for gene ontology (GO) terms related to synaptic transmission, ion transport, and cytoskeletal architecture. Statistical testing is detailed in Methods. **c.** Volcano plot showing effects sizes of aging on CpG methylation in the PWG cohort. TauSeverity feature CpGs are age-independent. **d.** Epigenetic age acceleration residuals (horizontal axis) calculated by the Horvath pan-tissue clock or Cortical brain-specific clock do not correlate with the actual hippocampal p-tau burden nor the age-adjusted p-tau residual. Statistical test was Pearson correlation. n = 260.

**Extended Data Fig. 4:**
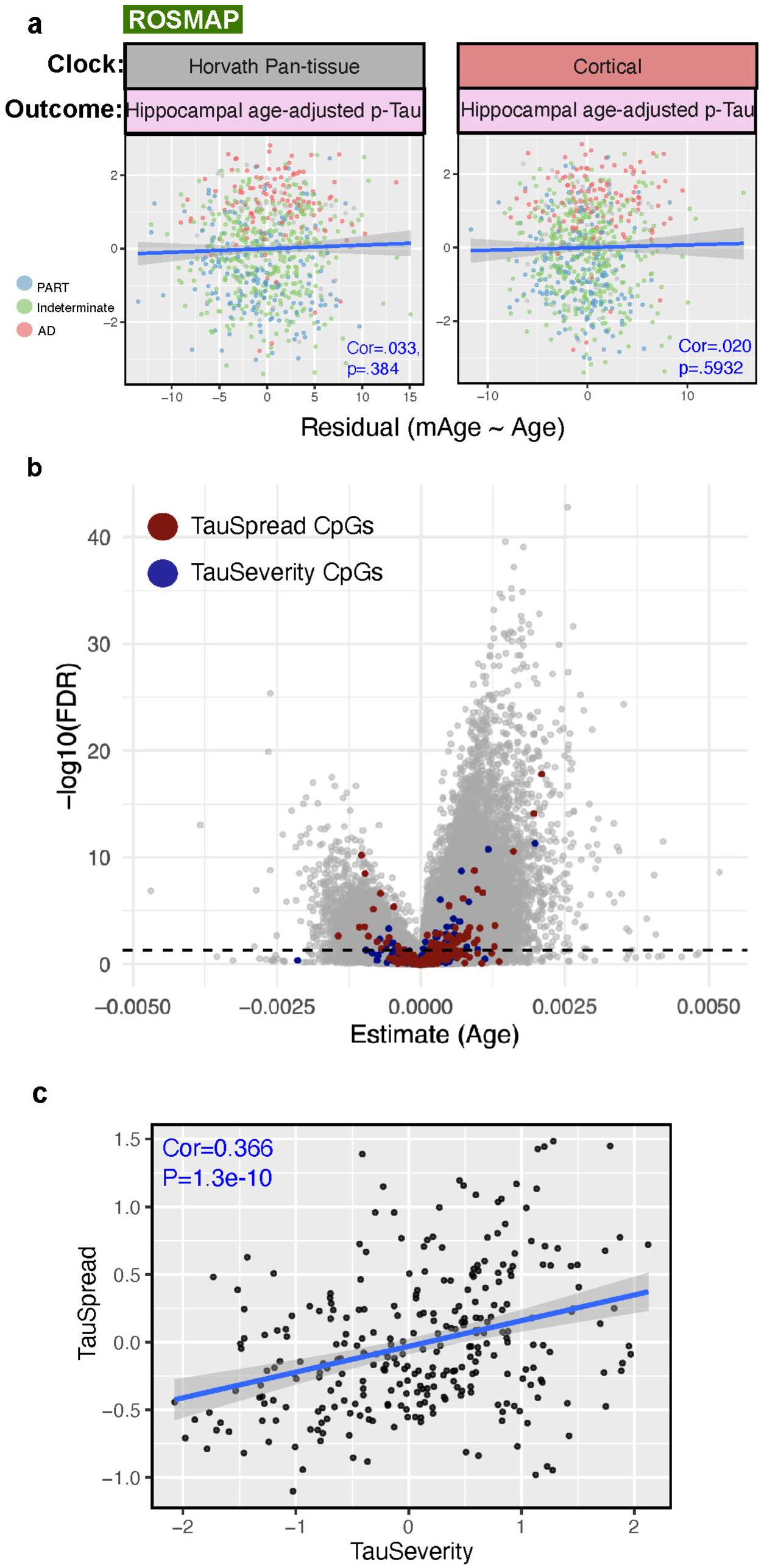
Characterization of ROSMAP TauSeverity and TauSpread CpGs. **a.** Epigenetic age acceleration (horizontal axis) calculated by the Horvath pan-tissue clock (left) and Cortical brain-specific clock (right) do not correlate with hippocampal age-adjusted p-tau residuals in the ROSMAP cohort. Statistical test was Pearson correlation: n = 707 [Horvath], 707 [Cortical]. **b.** Volcano plot demonstrating that TauSeverity (blue) and TauSpread (red) feature CpGs are largely age-independent in the ROSMAP cohort. **c.** TauSeverity and TauSpread are moderately correlated (Pearson’s cor = 0.366, P = 1.3e-10. One outlier is not displayed due to axes limits but was included in statistical testing. n = 291.

**Extended Data Fig. 5:**
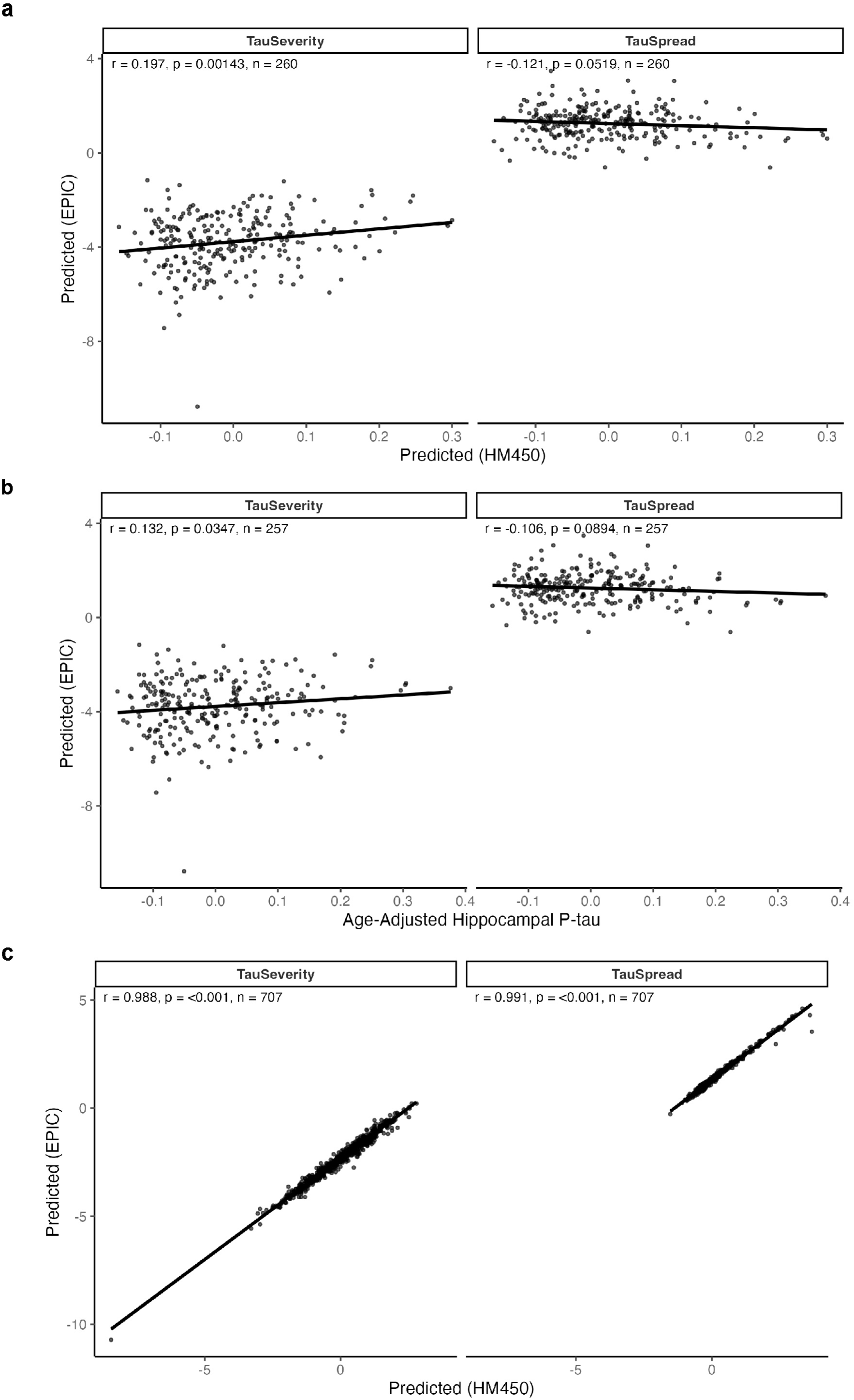
Cross-validation of ROSMAP TauSeverity in the PWG cohort. **a.** The PWG TauSeverity model predictions correlate with a ROSMAP TauSeverity model in which CpG features not present on the MethylationEpic BeadChip Array were excluded. **b.** The ROSMAP TauSeverity model in which CpG features not present on the MethylationEpic BeadChip Array were excluded predicts age-adjusted hippocampal p-tau residuals in external PWG cohort. **c.** Excluding CpGs represented on the HumanMethylation 450k (HM450) BeadChip Array but not on the MethylationEPIC BeadChip Array does not significantly alter the prediction of TauSeverity in the ROSMAP cohort.

**Extended Data Fig. 6:**
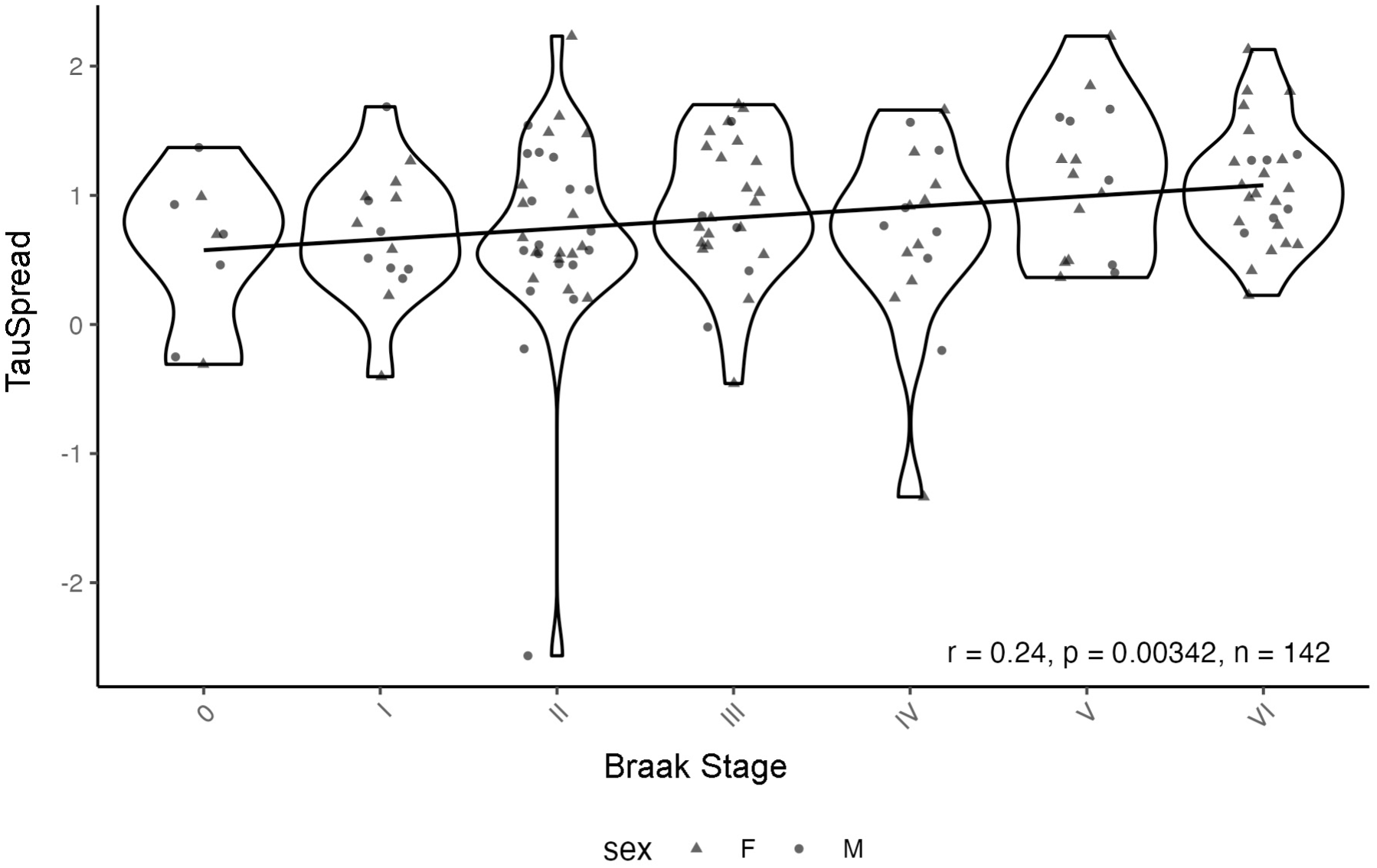
Cross-validation of ROSMAP TauSpread in the MSBB cohort. The ROSMAP TauSpread model when applied to frontal cortex DNAm from the external Mount Sinai Brain Bank (MSBB) cohort correlates with Braak NFT stage, an indicator of the anatomical spread of - p-tau pathology.

**Extended Data Fig. 7:**
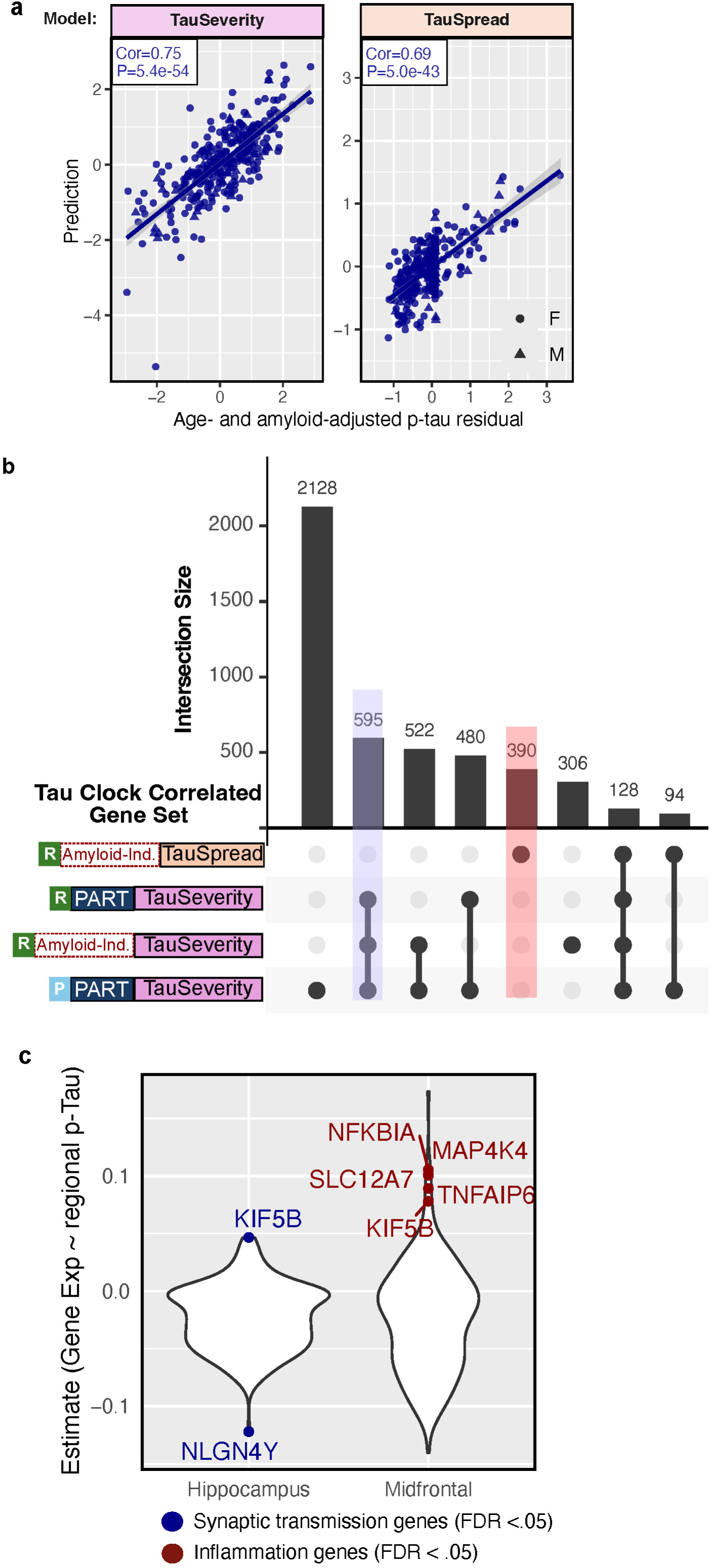
Identification of amyloid-independent CpG modifiers of p-tau and associated genes. **a.** Amyloid-adjusted TauSeverity and TauSpread respectively predict hippocampal (left) and midfrontal (right) age- and ß-amyloid-adjusted p-tau residuals with high accuracy in the ROSMAP cohort. [hippocampus, TauSeverity] n = 291, Pearson Cor = 0.75, P = 5.35e-54; [midfrontal cortex, TauSpread] n = 291, Pearson Cor = 0.69 and P = 5.02e-43. F = female, M = male. For TauSeverity one outlier is not displayed due to axes limits but was included in statistical testing. **b.** Upset plot demonstrating number of overlapping and distinct genes for different Tau clock model CpG sets. Genes correlated with methylation of the model CpGs were identified. “Amyloid-independent” corresponds to the CpG sets from Fig. 2C shared between amyloid and non-amyloid adjusted models. R = ROSMAP, P = PWG. Fig. is truncated to highlight overlap. Complete listing of genes in Table S3. **c.** Violin plots illustrating the effect size of p-tau severity in hippocampus or spread to midfrontal cortex on gene expression for synaptic transmission genes identified from TauSeverity models (left) and inflammation related genes from TauSpread models (right). Significant genes (FDR < .05) highlighted. Statistical testing is detailed in Methods.

**Extended Data Fig. 8:**
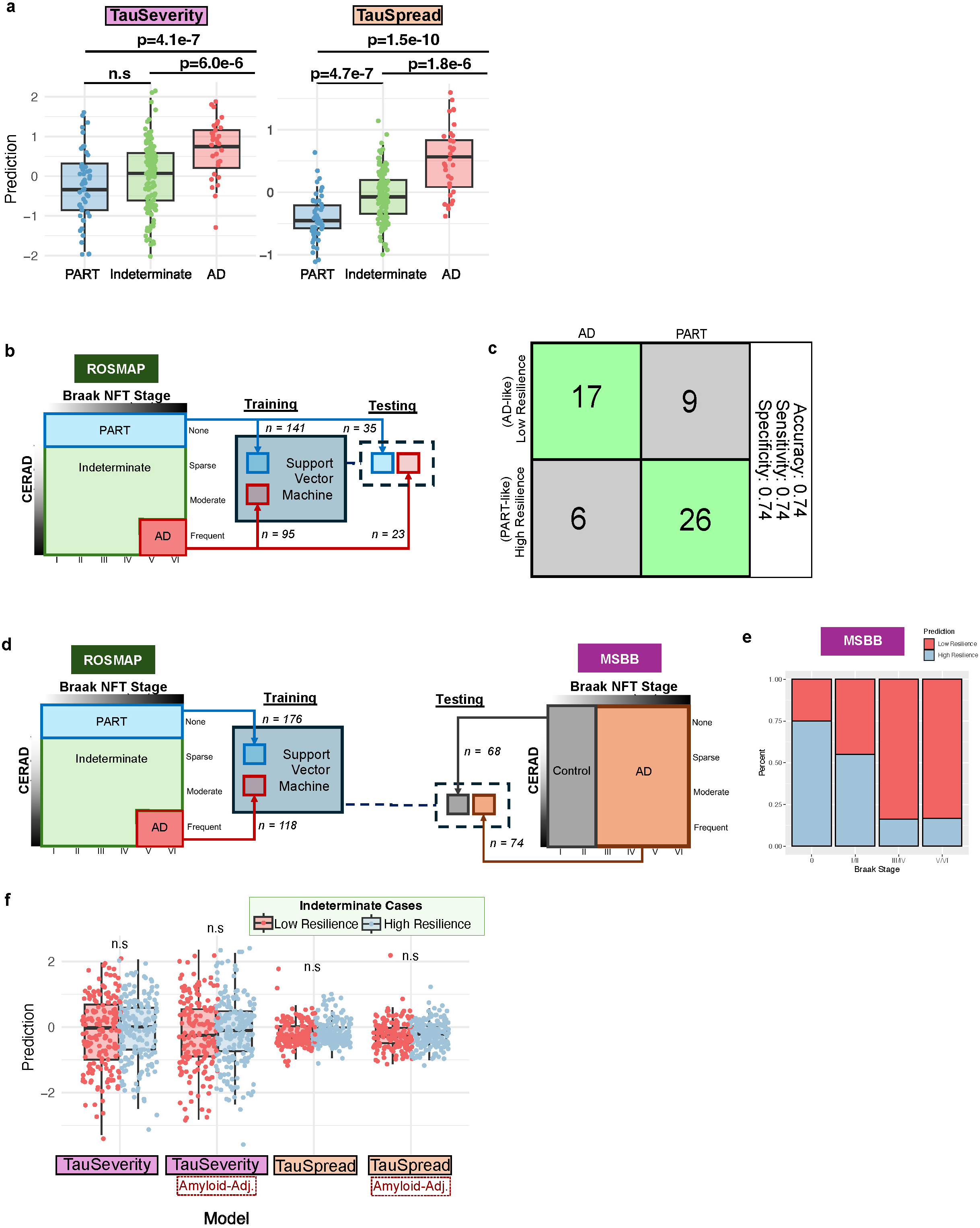
DNAm distinguishes Indeterminate cases along the PART-AD continuum. **a.** TauSeverity (left) and TauSpread (right) increase along the PART-AD continuum. TauSpread more strongly distinguishes Indeterminate cases from PART. For TauSeverity, one outlier is not displayed due to axes limits but was included in statistical testing. n= 290. **b.** Schematic of training and testing of the preliminary ResilienceDetector. DNAm from ROSMAP cases meeting definition for PART (n=141) or AD (n=95) were used to train a support vector machine, which was tested for accuracy on the remaining PART (n=35) or AD (n=23) cases in the ROSMAP cohort. **c.** Confusion matrix demonstrating performance of the preliminary ResilienceSwitch on testing cases. **d.** Schematic of training and testing of the final ResilienceSwitch. DNAm from all ROSMAP cases meeting definition for PART (n=176) or AD (n=118) were used to train a support vector machine, which was then applied to an external cohort from the Mount Sinai Brain Bank (MSBB). **e.** Proportion of cases by Braak NFT stage within the MSBB cohort that are classified as High or Low Resilience by the ResilienceDetector. Note that in this cohort, cases are defined as AD (Braak NFT stage III-VI) or control (Braak NFT stage 0-II), without regard to ß-amyloid status. Therefore, some cases that meet our definition of PART may be present in both groups. **f.** High and Low Resilience cases in the ROSMAP cohort could not be distinguished by either unadjusted or amyloid-adjusted Tau clock models. For TauSeverity, one outlier is not displayed due to axes limits but was included in statistical testing. Pairwise t-tests: n = 405. n.s. = not significant.

**Extended Data Fig. 9:**
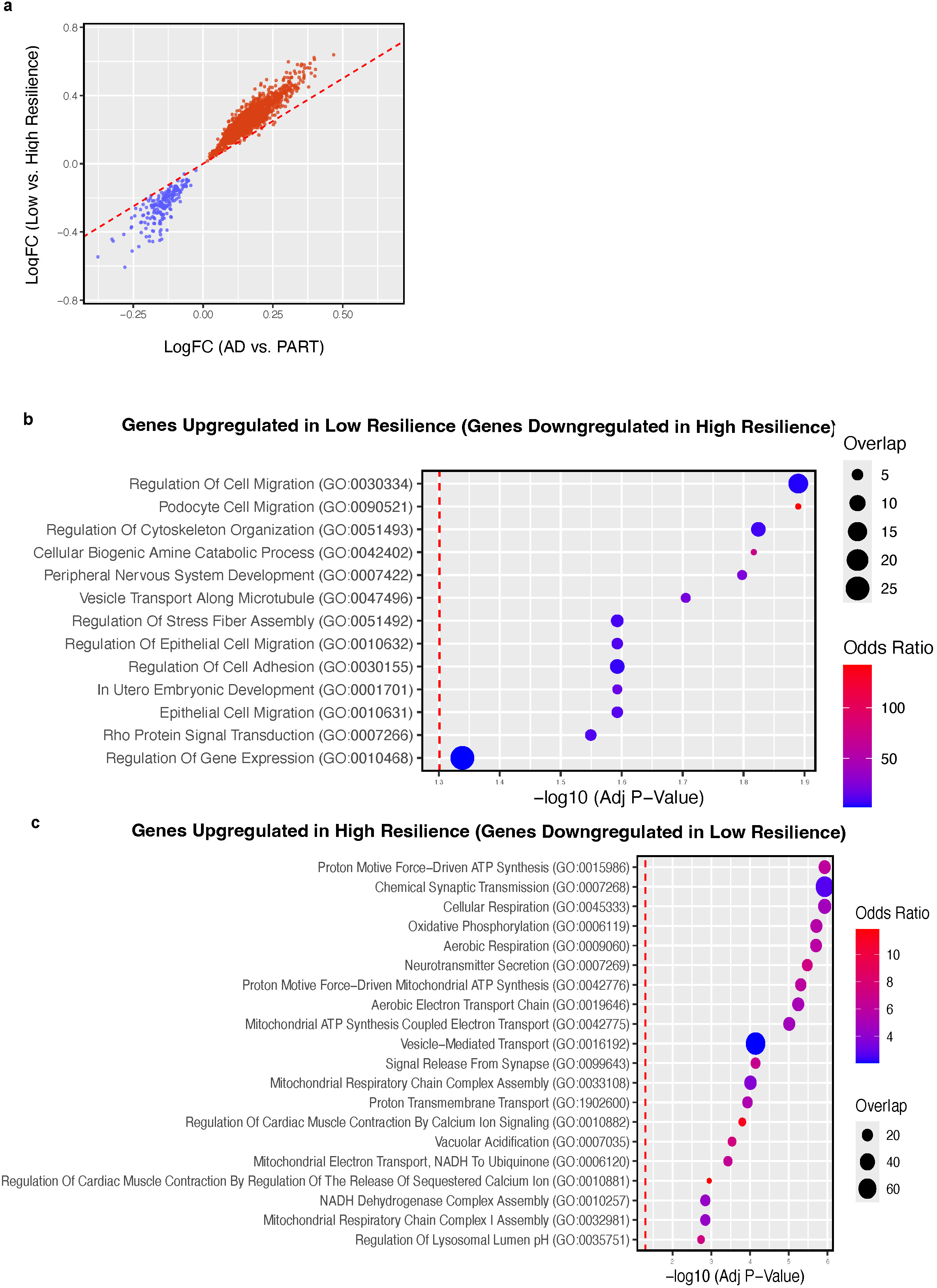
High and Low Resilience cases have a distinct transcriptomic signature. **a.** Correlation of differential gene expression effect size between AD vs. PART cases (horizontal axis) and Low vs. High Resilience cases (vertical axis) for all highlighted genes in Fig. 3D. Pearson Cor = 0.97 and P < 2.2e-16. **b.** Genes upregulated Low versus High Resilience cases are enriched for gene ontology (GO) terms related to cell differentiation and migration. Statistical testing is detailed in Methods. **c.** Genes upregulated in High compared to Low Resilience cases are enriched for gene ontology (GO) terms related to oxidative phosphorylation and cellular respiration. Statistical testing is detailed in Methods.

**Supplemental Table 1:**
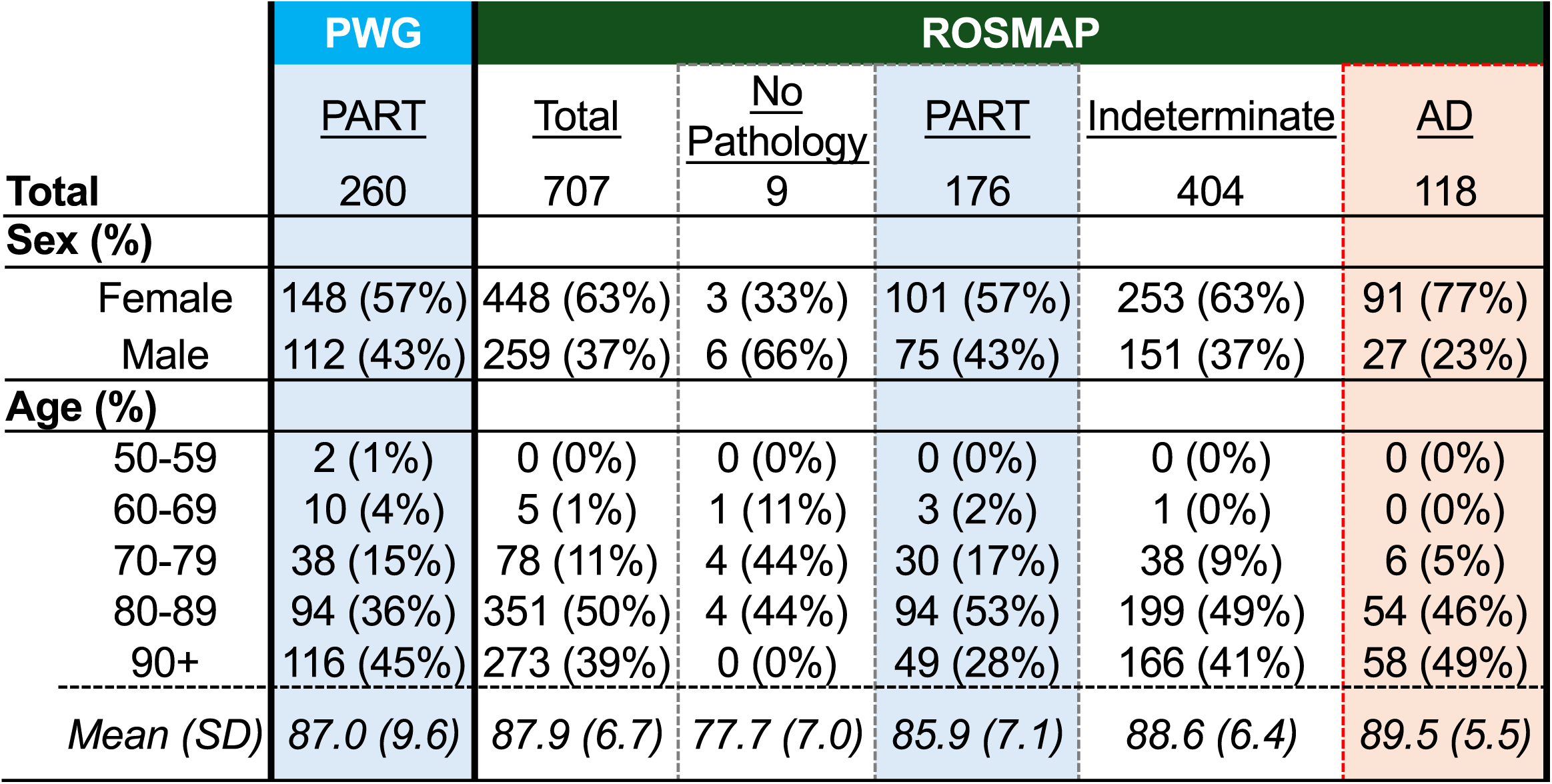
Individual characteristics for the PART Working Group (PWG) and Religious Orders Study and Memory and Aging Project (ROSMAP) cohorts separated by neuropathological case definitions (Methods).

## Appendix

The PART working group: Jean-Paul Vonsattel (Department of Pathology and Cell Biology, Department of Neurology, and the Taub Institute for Research on Alzheimer’s Disease and the Aging Brain, Columbia University Medical Center, New York, NY), Andy F. Teich (Department of Pathology and Cell Biology, Department of Neurology, and the Taub Institute for Research on Alzheimer’s Disease and the Aging Brain, Columbia University Medical Center, New York, NY), Marla Gearing (Department of Pathology and Laboratory Medicine (Neuropathology) and Neurology, Emory University School of Medicine, Atlanta, GA), Jonathan Glass (Department of Pathology and Laboratory Medicine (Neuropathology) and Neurology, Emory University School of Medicine, Atlanta, GA), Juan C. Troncoso (Department of Pathology, Division of Neuropathology, Johns Hopkins University School of Medicine, Baltimore, MD), Matthew P. Frosch (Department of Neurology and Pathology, Harvard Medical School and Massachusetts General Hospital, Charlestown, MA), Bradley T. Hyman (Department of Neurology and Pathology, Harvard Medical School and Massachusetts General Hospital, Charlestown, MA), Melissa E. Murray (Department of Neuroscience, Mayo Clinic, Jacksonville, FL), Johannes Attems (Translational and Clinical Research Institute, Newcastle University, Newcastle upon Tyne, UK), Margaret E. Flanagan (Department of Pathology (Neuropathology), Northwestern Cognitive Neurology and Alzheimer Disease Center, Northwestern University Feinberg School of Medicine, Chicago, IL), Qinwen Mao (Department of Pathology (Neuropathology), Northwestern Cognitive Neurology and Alzheimer Disease Center, Northwestern University Feinberg School of Medicine, Chicago, IL), M-Marsel Mesulam (Department of Pathology (Neuropathology), Northwestern Cognitive Neurology and Alzheimer Disease Center, Northwestern University Feinberg School of Medicine, Chicago, IL), Sandra Weintraub (Department of Pathology (Neuropathology), Northwestern Cognitive Neurology and Alzheimer Disease Center, Northwestern University Feinberg School of Medicine, Chicago, IL), Randy L. Woltjer (Department of Pathology, Oregon Health Sciences University, Portland, OR), Thao Pham (Department of Pathology, Oregon Health Sciences University, Portland, OR), Julia Kofler (Department of Pathology [Neuropathology], University of Pittsburgh Medical Center, Pittsburgh, PA), Julie A. Schneider (Departments of Pathology [Neuropathology] and Neurological Sciences, Rush University Medical Center, Chicago, IL), Lei Yu (Departments of Pathology [Neuropathology] and Neurological Sciences, Rush University Medical Center, Chicago, IL), Dushyant P. Purohit (Department of Pathology, Neuropathology Brain Bank and Research CoRE, Icahn School of Medicine at Mount Sinai, James J. Peters VA Medical Center, New York, NY), Vahram Haroutunian (Department of Psychiatry, Alzheimer’s Disease Research Center, James J. Peters VA Medical Center, Nash Department of Neuroscience, Ronald M. Loeb Center for Alzheimer’s Disease, Friedman Brain Institute, Icahn School of Medicine at Mount Sinai, New York, NY), Patrick R. Hof (Nash Department of Neuroscience, Ronald M. Loeb Center for Alzheimer’s Disease, Friedman Brain Institute, Icahn School of Medicine at Mount Sinai, New York, NY), Sam Gandy (Departments of Psychiatry and Neurology, Center for Cognitive Health, Alzheimer’s Disease Research Center, James J. Peters VA Medical Center, Icahn School of Medicine at Mount Sinai, New York, NY and Department of Icahn School of Medicine at Mount Sinai, New York, NY), Mary Sano (Department of Psychiatry, Alzheimer’s Disease Research Center, James J. Peters VA Medical Center, Icahn School of Medicine at Mount Sinai, New York, NY), Thomas G. Beach (Department of Neuropathology, Banner Sun Health Research Institute, Sun City, AZ), Wayne Poon (Department of Neurology, Department of Epidemiology, Institute for Memory Impairments and Neurological Disorders, UC Irvine, Irvine, CA), Claudia H. Kawas (Department of Neurology, Department of Neurobiology and Behavior, Institute for Memory Impairments and Neurological Disorders, UC Irvine, Irvine, CA), María M. Corrada (Department of Neurology, Department of Epidemiology, Institute for Memory Impairments and Neurological Disorders, UC Irvine, Irvine, CA), Robert A. Rissman (Department of Neurosciences University of California and the Veterans Affairs San Diego Healthcare System, La Jolla, San Diego, CA), Jeff Metcalf (Department of Neurosciences University of California and the Veterans Affairs San Diego Healthcare System, La Jolla, San Diego, CA), Sara Shuldberg (Department of Neurosciences University of California and the Veterans Affairs San Diego Healthcare System, La Jolla, San Diego, CA), Bahar Salehi (Department of Neurosciences University of California and the Veterans Affairs San Diego Healthcare System, La Jolla, San Diego, CA), Peter T. Nelson (Department of Pathology [Neuropathology] and Sanders-Brown Center on Aging, University of Kentucky, Lexington, KY), Edward B. Lee (Center for Neurodegenerative Disease Research, Department of Pathology and Laboratory Medicine, Perelman School of Medicine, University of Pennsylvania, Philadelphia, PA), David A. Wolk (Department of Neurology, Perelman School of Medicine, University of Pennsylvania, Philadelphia, PA), Corey T. McMillan (Department of Neurology, Perelman School of Medicine, University of Pennsylvania, Philadelphia, PA), C. Dirk Keene (Department of Laboratory Medicine and Pathology, University of Medicine, Seattle, WA), Caitlin S. Latimer (Department of Laboratory Medicine and Pathology, University of Medicine, Seattle, WA), Thomas J. Montine (Department of Laboratory Medicine and Pathology, University of Medicine, Seattle, WA and Department of Pathology, Stanford University, Palo Alto, CA), Gabor G. Kovacs (Laboratory Medicine Program, Krembil Brain Institute, University Health Network, Toronto, ON, Canada, Tanz Centre for Research in Neurodegenerative Disease and Department of Laboratory Medicine and Pathobiology, University of Toronto, Toronto, Ontario, Canada, and Institute of Neurology, Medical University of Vienna, Vienna, Austria), Mirjam I. Lutz (Institute of Neurology, Medical University of Vienna, Vienna, Austria), Peter Fischer (Department of Psychiatry, Danube Hospital, Vienna, Austria), Richard J Perrin (Department of Pathology and Immunology, Department of Neurology, Knight Alzheimer Disease Research Center, Washington University School of Medicine, St. Louis, MO), Nigel J. Cairns (College of Medicine and Health, University of Exeter, Exeter, UK).

